# Airborne Pathogen Detection in Fine Aerosol Exhaled Breath Condensates

**DOI:** 10.1101/2022.05.25.22275435

**Authors:** John Henderson, Theodora Mantso, Saqib Ali, Rüdiger Groß, Janis A. Müller, Amie Wilkinson, Kavit Shah, Louise Usher, Beth Auld, Andrew Nelson, William Cheung, Anil Namdeo, Madeleine Combrinck, Phil Hackney, Volkan Turgul, Edison Jahaj, Nikolaos Athanasiou, Taxiarchis Nikolouzakis, Pedro J. Almeida, Chrysa Rokka, Daniel C. Queiroz, Edward Wright, Alexandros Zafiropoulos, Izzet Kale, Darren Smith, Diamantis P. Kofteridis, Aristides Tsatsakis, Jan Münch, Paraskevi A. Katsaounou, Anastasia Kotanidou, Pagona Lagiou, Gkikas Magiorkinis, Renato S Aquiar, Mauro M. Teixeira, Sterghios A. Moschos

## Abstract

**Rationale:** Exhaled breath condensate (EBC) promises a valuable, non-invasive, and easy to obtain clinical sample. However, it’s not currently used diagnostically due to poor reproducibility, sample contamination, and sample loss.

**Objective:** We evaluated whether a new, hand-held EBC collector (PBM-HALE^TM^) that separates inertially impacted large droplets (LD) before condensing fine aerosols (FA) in distinct, self-sealing containers, overcomes current limitations.

**Methods:** Sampling consistency was determined in healthy volunteers by microbial culture, 16S phylogenetics, spectrophotometry, RT-PCR, and HILIC-MS. Capture of aerosolised polystyrene beads, liposomes, virus-like particles, or pseudotyped virus was analysed by nanoparticle tracking analysis, reporter expression assays, and flow cytometry. Acute symptomatic COVID-19 case tidal FA EBC viral load was quantified by RT-qPCR. Exhaled particles were counted by laser light scattering.

**Measurements and Main Results:** Salivary amylase-free FA EBC capture was linear (R^2^=0.9992; 0.25-30 min) yielding RNA (6.03 μg/mL) containing eukaryotic 18S rRNA (RT-qPCR; p<0.001) but not human GAPDH, RNase P, or beta actin mRNA;141 non-volatile metabolites included eukaryotic cell membrane components, and cuscohygrine 3 days after cocaine abuse. Culturable aerobe viability was condensation temperature-dependent. Breath fraction-specific microbiota were stable, identifying *Streptococcus* enrichment in a mild dry cough case. Nebulized pseudotyped virus infectivity loss <67% depended on condensation temperature, and particle charge-driven aggregation. SARS-CoV-2 RNA genomes were detected only by forced expiration FA EBC capture, in 100% of acute COVID-19 patients.

**Conclusions:** High purity, distal airway FA EBC can reproducibly and robustly inform contamination-free infectious agent emission sources, and be quantitatively assayed for multiple host, microbial, and lifestyle biomarker classes.

## Introduction

Air expelled from the lungs becomes humidified by water vapour and fluid lining the respiratory tract epithelium, as well as the oral and nasal epithelium, by means of the ‘fluid film burst’ mechanism (1) resulting in aerosols released as a component of exhaled breath. These dry and aqueous particles are predominantly fine aerosol (FA) particles <10 μm in diameter (2), as well as large droplets (LD). Fine aerosols originate from distal airways and alveoli, while most large droplet production is physically restricted to the upper airways and oropharyngeal epithelium (3).

Upon exhalation, both particle types can remain airborne (4), depending on air movement, ambient temperature, and relative humidity (5) since these physical parameters drive particle size evolution: hydration-driven swelling leads to gravitational sedimentation, whereas evaporative shrinking drives extended airborne diffusion (6). Consequently, selective capture and analysis of exhaled FA offers the unique opportunity to specifically analyse metabolites, pathogens, and biomolecules originating from the distal lung with minimal upper respiratory or oral content cross-contamination. Such a solution could potentially substitute invasive methods e.g. tracheal aspirates and bronchoalveolar lavage, or potentially infectious aerosol-generating sputum induction, methods which inherently suffer sample cross-contamination from the upper respiratory tract.

Cooling to condense exhaled breath (7–11) yields a condensate of respiratory gases, aerosols, and droplets collectively known as exhaled breath condensate (EBC). The resulting aqueous sample can be analysed by ELISA, western blotting, mass spectrometry, PCR, RT-PCR, and HPLC (3–7). Differential levels of cytokines, growth hormones, lipids, microRNAs, and distinct metabolomic profiles, as well as volatile compound signatures, have been correlated with pathological states including lung cancer (8,9), pulmonary fibrosis (10), bronchoconstriction (11), physiological shock (12) and even neurological disorders (12).

Despite the apparent diagnostic potential of EBC, clinical adoption has been restricted to volatile compound mass spectrometry for the detection of *Helicobacter pylori* gastric infection (13). Challenges beyond this indication centre on the poor reliability of EBC collectors (3, 14); thus, in 2005 (14) and again in 2017 (3), Horvàth *et al.* highlighted key technical issues regarding EBC collection, and the need to establish consistent practices for collection and analysis. Yet until 2019, despite the substantial unmet need for diagnosing acute respiratory infections among the elderly infirm (15), the nature of pediatric wheeze (16), or indeed the persistent threat of aerosolized bioweapons (17), limited progress had been made in solving these problems. Instead, breath diagnostics focused on volatile compound analysis (18–22) due to the commercial facilitation offered from the regulatory approval of *H. pylori* breath testing in the USA.

To specifically address the key technical challenges of EBC sampling reproducibility, salivary and ambient matter contamination, sample loss, and healthcare professional safety (3, 14), we have developed a new, hand-held exhaled breath condensate collector (PBM-HALE^TM^). This device inertially impacts and collects the LD fraction of exhaled breath, before condensing FA with enhanced efficiency in a separate specimen container. The aims of this study were to evaluate the capacity of this device to isolate distal lung FA EBC free of oral and upper airway contaminants, the classes of biomarkers identifiable in the resulting FA EBC samples, and whether putative causal agents of symptomatic respiratory infections can be safely collected and detected in orally exhaled tidal breath.

## Methods

### Participant Recruitment

All healthy volunteer EBC samples used within this study were obtained with participant informed consent under Northumbria University ethics application no. 43341 approved by the Department of Applied Sciences Subcommittee of the University Research Ethics Committee. All COVID-19 patient EBC samples were collected with informed consent under the National and Kapodistrian University of Athens General Hospital ‘Evangelismos’ ethics application protocol no. 280/24-4-2020 approved by the Scientific Committee of the General Hospital ‘Evangelismos’ and the approval no. 54358021.1.0000.5149 by the Institutional Review Board of the Federal University of Minas Gerais. Patients in Greece were recruited among attendees of the Emergency Department of Evangelismos Hospital, Athens, Greece, and the University General Hospital of Herakleion, Herakleion, Crete, Greece between June 2020 and June 2022. Patients in Brazil were recruited among attendees of the suburban primary care centre Centro de Saúde Jardim Montanhes, Center for Advanced and Innovative Therapies, Federal University of Minas Gerais, Minas Gerais, Belo Horizonte, Brazil, between March and July 2022. Participants were included in the study if acutely symptomatic (days 0-5) for COVID-19 (fever, persistent cough, dysgeusia or dysosmia, or dyspnoea) and confirmed positive for SARS-CoV-2 by nasopharyngeal swab lateral flow test. Proper nasopharyngeal specimen collection from a trained medical doctor was performed according to CDC guidelines using sterile mini tip swabs with a flexible plastic shaft. As soon as the patient’s head was tilted back 70°, the swab was inserted slowly through the nostril parallel to the palate until resistance was encountered, then rubbed, rolled, and left in place for several seconds to absorb secretions. Finally, the swab was removed while rotating it and placed into a vial containing viral transport media. Additional samples were obtained from cases at days 0-5 from admission or convalescents within COVID-19 treatment wards.

### EBC Sampling Devices

Early experiments were carried out using 50 mL Corning^®^ preassembled closed system solution centrifuge tubes as the FA condensing surface and EBC vessel, connected to 5mm internal diameter, 3cm length Teflon-coated tubing fitted with pinch cock tube clamps (Thermo Fisher Scientific, Loughborough, UK), and a square saliva trap mouthpiece (Alcopro, Knoxville, TN) as the LD inertial impaction and vessel (Figure 1). The condensate collector lid was modified internally in line with Dreschel bottle principles to lengthen the fluid travel path and maximise breath contact with the condensing surface prior to breath egress via a second Teflon-coated tube. Clamps were used to prevent environmental contamination and sample loss before and after sampling. Condensation temperature control was provided by means of a custom-built centrifuge tube holder (room temperature (RT) condensation), or polysterene coolbox immersion in wet (0°C) or dry ice (−78.5°C).

**Figure 1:**
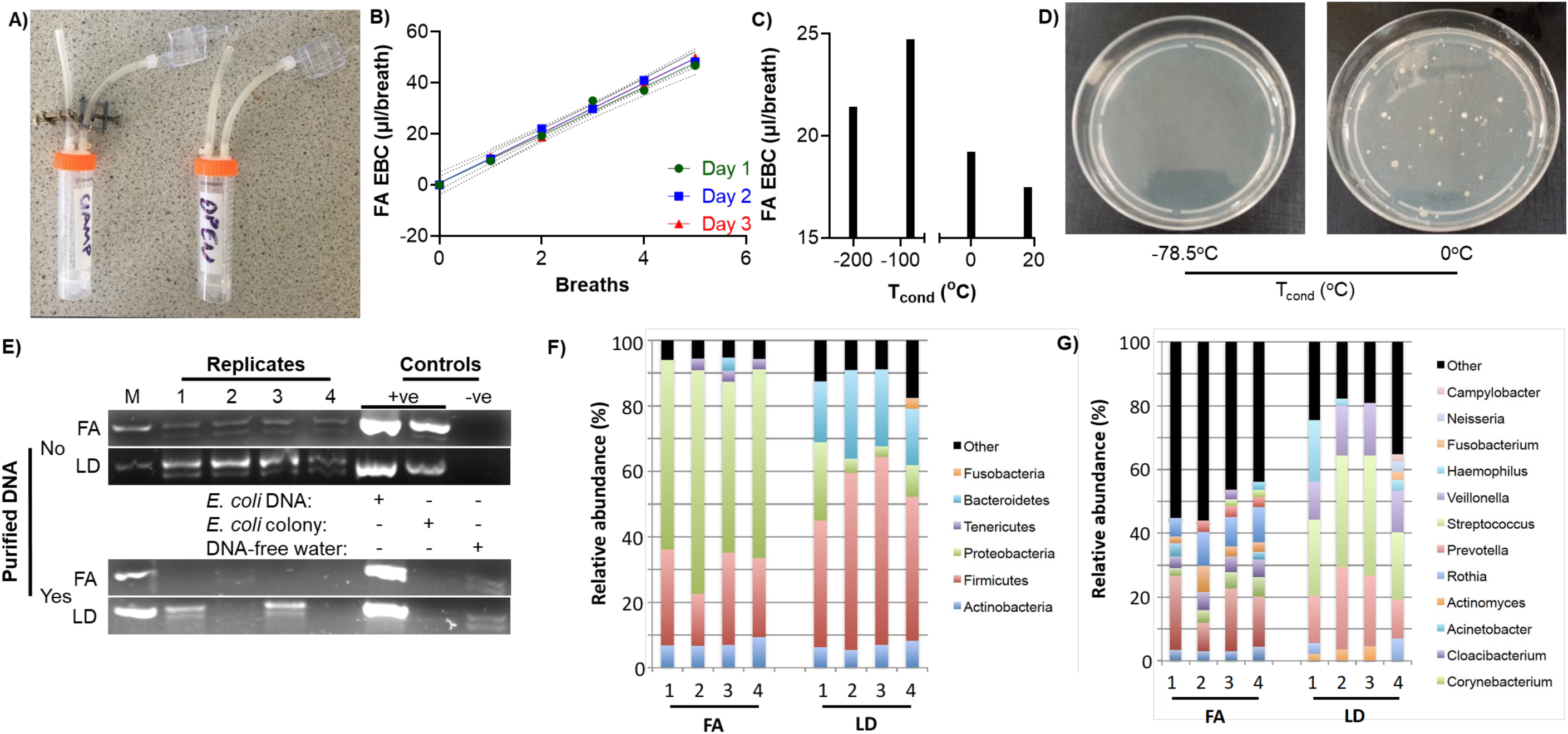
FA EBC consistency in an early laboratory prototype impactor/condenser breath collector. (A) This assembly involves a minimal distance from the LD-trapping inertial impactor saliva trap to the FA EBC collector to eliminate condensate formation in conducting tube walls and focus condensation of the terminal volume of the exhalation column during the inspiration phase. The thumbscrew clamps prevent environmental contamination or sample evaporation when the sampler is not in use. (B) The volume of FA EBC captured was determined as a function of the number of exhalations of a healthy adult female volunteer sampled over 3 consecutive days (n=3 per day, 95% CI shown). (C) The condensation temperature-dependent FA EBC collection rate was calculated from the slope of EBC volume per number of breath experiments (n=3 per condensation temperature) conducted with one healthy adult female volunteer on the same day. (D) The viability of bacteria in the FA EBC as a function of condensation temperature was determined by nutrient agar plating the full volume produced with a 5-breath sample (n=3, representative plates shown). (E) 16S rRNA gene V3 region DNA amplicons were generated with KAPA Plant 3G polymerase either directly from LD or EBC FA obtained under −78.5°C condensation, or from purified DNA from the same samples. ‘M’: 100 bp marker; ‘DNA’ and ‘colony’ controls correspond to *Escherichia coli* DNA extract and colony PCR. Negative control water reactions consisted of undiluted PCR mastermix supplemented with 16S rRNA V3 primers only. (F, G) Phylum-(F) and genus-level (G) microbiomic profiling of FA and LD samples generated by 10 tidal oral exhalations, obtained over 4 consecutive days from one adult healthy male volunteer. The samples were subjected to direct DNA amplification for the V3 region of the 16S rRNA gene using KAPA Plant 3G polymerase without prior DNA extraction/purification. OTUs with <1% relative abundance were grouped as “other” in F and G.

PBM-HALE^TM^ devices (Figure 2) and custom components were constructed in Solidworks v. 2016-2021sp3 (Dassault Systèmes, Corp., Waltham, MA), to be subsequently prototyped and produced by additive manufacturing using a Stratasys Objet 30 3D printer, Objet 3D Fullcure 705 support resin and Objet VeroWhitePlus FullCure 835 (Tri-tech 3D Ltd., Stoke on Trent, UK). The coolant chamber was manufactured using a Prusa I3 MK3 3D printer and Prusament PLA vanilla white filament (Prusa Research, Prague, Czech Republic) with 50mm grey Craftfoam (Panel Systems Ltd., Sheffield, UK) used as coolant chamber insulation.

**Figure 2:**
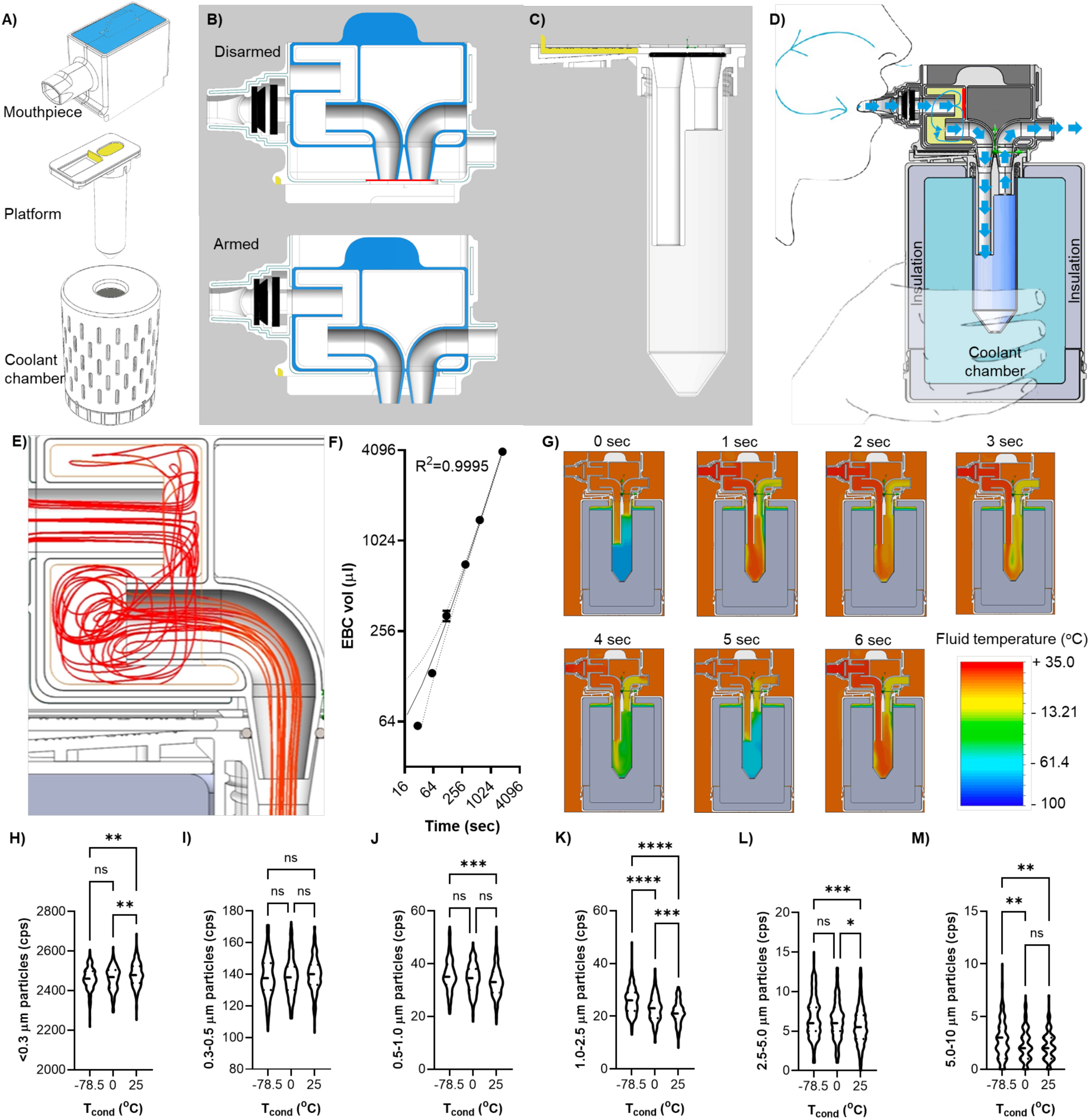
The PBM-HALE^TM^ EBC collector linearly captures a FA EBC sample by inducing aerosol swelling when flow is paused through the device during inhalation. (A) The components of the 3D printed PBM-HALE^TM^ device (see video 1 online for assembly sequence). The 50mL FA EBC condensing vial (platform) is inserted into the coolant chamber which is then loaded with coolant. The mouthpiece is then racked onto the platform in a right-to-left motion. The platform lid features a drawer (yellow) which is retracted to expose the conducting tubes inside the FA EBC condensing vial, only when the mouthpiece is mounted on the assembly. (B) Section cut of the mouthpiece in the armed and disarmed configurations. Mounting the mouthpiece onto the platform retracts the platform drawer by catching the drawer upward facing lip seen in (A) in a notch marked in (B) in yellow, situated at the underside of the mouthpiece, thus exposing the conducting tubes in the FA EBC condensing vial. The mouthpiece has a sliding inner body (light blue in A, B) that prevents the mouthpiece’s conducting tubes from being contaminated before and after use. Pressing down on the mouthpiece inner body (arming) brings its’ internal tubing into fluid communication with the inlet and outlet tubes of the outer body. The motion also breaks a protective foil seal (red line) at the bottom of the outer body, bringing the conical connectors into fluid communication with the conducting tubes inside the FA EBC condensing vial. (C) Section cut of the platform component showing the architecture of the platform lid and FA EBC condensing vial. The drawer (yellow) in the lid of the platform is shown in the retracted position. The mouthpiece inner body conical connectors interface with the platform lid conducting tubes upon mouthpiece arming, forming a seal with two O rings (black). (D) The internal architecture of the assembled device in the armed position during use. A one-way valve (black; seen in B and D) prevents inhalation through the device. The cavity (yellow) inside the inner body of the mouthpiece contains an ‘S’ bend architecture presenting a perpendicular impaction surface (red line) to the breath (solid blue arrows) within 3 cm of the breath inlet. This creates turbulent flow (light blue arrows) within the inner mouthpiece cavity, enhancing LD elimination. The breath column proceeds into the FA EBC vial (gradient blue) which is immersed in coolant (teal). The exhalant exits via the elbow joint outlet. Upon sampling completion the mouthpiece is disarmed, unracked (simultaneously sealing the FA EBC condensing vial), and the FA EBC vial can be removed to storage. (E) Computational flow dynamics model of turbulent airflow (red lines) inside the mouthpiece inner body during tidal exhalation. (F) FA EBC volume capture as a log_2_-log_2_ function of time from a healthy adult female volunteer; dotted lines represent 95% CI. (G) One second interval heatmap of exhaled breath temperature within PBM-HALE^TM^ condensing at −78.5°C, during a 5sec, 0.5L tidal breath as modelled by computational flow dynamics (10,000 iteration convergence), starting at 0sec with the device pre-equilibrated to the coolant. (H-M) Particle counts at the PBM-HALE^TM^ outlet point as a function of condensation temperature (T_cond_) over a 3 min sampling period for an adult healthy female. Counts presented in interval bins of (H) <0.3 μm, (I) 0.3-0.5 μm, (J) 0.5-1.0 μm, (K) 1.0-2.5 μm, (L) 2.5-5.0 μm, and (M) 5-10.0 μm, with violin plots showing medians and quartiles; *: p<0.05; **: p<0.01; ***: p<0.001; ****: p<0.000; ns: not significant.

Working inside a class II microbial safety cabinet devices were decontaminated from analytes by immersion in a 0.6% sodium hypochlorite solution (Merck, Manchester, UK), rinsed with RNAse-free water (Thermo Fisher Scientific), air dried and assembled, before packing into sodium hypochlorite-decontaminated zip lock bags ahead of use. Unless otherwise stated, 50mL polypropylene Corning^®^ tubes were used as the FA EBC sample vessel using CentriStar^TM^ caps (high density polyethylene) or PBM-HALE^TM^ custom-built platform lids.

### Physical Parameter Measurement

Device temperature testing was performed using a Mastech MS6514 probe thermometer fitted with a type T thermocouple (Amazon, London, UK). Airborne particle size measurements were performed using a Particles Plus® 8506 Handheld Particle Counter fitted with a Particles Plus® temperature and humidity probe (Particles Plus, Inc., Stoughton, MA). Sample volumes were quantified using Sartorius Picus electronic single channel pipettes (Sartorius UK Ltd., Epsom, UK).

### Computational Flow Modelling

Computational flow and thermal modelling was executed in Soliworks v. 2021sp3 using a tidal breath flow model of 95% relative humidity exhaled breath at 35°C, 0.5L/3sec laminar flow. The duty cycle model employed was based on Matamedi-Fakr et al. (23) and Jawde et al. (24) with a 5sec period; expiration was modelled at 0.2L/sec, 0.15l/sec, 0.15L/sec flow for each of the first 3 seconds of each breathing cycle, followed by 0L/sec flow for the 2sec inspiration phase reflecting the inhalation prevention valve function in PBM-HALE^TM^. Flow and temperature calculations were computed for exhalation periods of 2min, 5min, and 15min of use against ambient conditions set at 20°C, 70% ambient relative humidity, 1Atm. Custom solid material models were defined as SYS Vero White (1175kg/m^3^, isotropic conductivity, absorption coefficient 0L/mm, 1.5 refractive index, 53°C melting temperature in line with the manufacturer product datasheet), and dry ice (1600kg/m^3^, specific heat of 1188 J/kg*K, isotropic, thermal conductivity in solid form of 0.2W/(m*K) based on previous reports (25, 26). External walls were set as adiabatic with whitewall default radiative surface and a 0.014mm roughness based on the 0.028mm layer thickness for the Objet printer assuming uniform layer contours.

### FA EBC, LD, and Saliva Sample Collection

All samples collected in this study were obtained by tidal oral breathing for the indicated time, or by singing as loudly as possible “happy birthday” for up to 15min. Unless otherwise stated, the PBM-HALE^TM^ cooling chamber was filled with powdered dry ice freshly produced using a CO_2_ cylinder (BOC Gas and Gear, Birtley, UK) fitted with a Bel-Art^TM^ SP Scienceware^TM^ Frigimat^TM^ Junior Dry Ice Maker (Thermo Fisher Scientific) immediately prior to sample collection. Alternatively, commercially procured dry ice pellets (∼1×3cm) or crushed wet ice was used. Dry ice was replenished every 30 minutes during sampling.

The FA EBC sample container was exposed to the environment just before initiating sampling and immediately isolated upon sampling completion. For the early prototype experiments this involved opening and closing the pinch cock tube clamps. For PBM-HALE^TM^, the mouthpiece was locked onto the coolant chamber and the inner mouthpiece part actuated into the armed position only immediately prior to initiating sampling (Figure 2 and video 1). After sample collection, the inner mouthpiece was returned to the unarmed position, and the mouthpiece was unloaded; two motions that physically isolate the LD within the mouthpiece, and return the custom platform lid into the closed position isolating the FA EBC. Samples were placed on wet ice before centrifuging at 4,000xg for 1 minute to pool the EBC at the bottom of the tube. The sample was then either immediately processed or stored at −80°C. The LD fraction was removed from PBM-HALE^TM^ by syringe and needle puncture of the saliva trap-containing mouthpiece. Fresh saliva samples were collected by drooling for 2 minutes into a microcentrifuge tube, spun at 4,000xg for 1 minute, after which the supernatant was added to a clean tube to separate it from any oral debris.

### Protein analysis

Sample volumes of 0.18mL were added to 0.02mL of 10x RIPA buffer (100mM Tris-Cl pH 8.0, 0.5% sodium deoxycholate, 0.5% SDS, 10% Triton X-100, 1.4M NaCl; Thermo Fisher Scientific), chilled on ice for 25min and vortexed at maximum speed intermittently. Lysate volumes of 0.15mL and bovine serum albumin protein standards (2 – 20ng; Thermo Fisher Scientific) were then mixed with 0.15mL of the micro BCA kit working reagent (Thermo Fisher Scientific) and absorbance was measured at 562nm after incubating at 37°C for 4-6 hours using a Spark® Cyto 96 well plate reader (Tecan, Männedorf, Switzerland). Sample protein concentrations were then calculated using the equation of the line of best fit for the standard curve. Undiluted tidal breath LD samples, or fresh saliva samples subjected to a freeze-thaw cycle on dry ice and diluted 1:200 with physiological saline were analysed using an α-amylase kinetic assay (Salimetrics LLC, Carlsbad, CA) following the manufacturer’s protocol.

### RT-PCR

For laboratory samples, RNA was extracted by adding 0.2mL of each sample to 0.8mL of TRIzol (Thermo Fisher Scientific) and virgorously vortexing for 1min. An 80% fraction of the upper aqueous phase was removed to prevent interphase DNA contamination. After precipitating in 100% isopropanol and washing with 75% ethanol (Thermo Fisher Scientific), RNA was eluted in 0.02mL nuclease-free water. RNA concentrations were then quantified using an ND-1000 Nanodrop spectrophotometer (Thermo Fisher Scientific) at an absorbance of 260nm. RNA samples were converted to cDNA using a maxima first strand cDNA synthesis kit (Thermo Fisher Scientific). qPCR was performed using a PrecisionPLUS qPCR mastermix with SYBR Green (PrimerDesign Ltd., Chandler’s Ford, UK) using either pan-eukaryotic 18S rRNA, human GAPDH, or human β-actin primers (Eurofins Scientific, Luxembourg). Thermal cycling was performed on a BioRad CFX96 (BioRad, Watford, UK) as follows: 2min at 95°C, then 40 cycles of 10sec at 95°C and 60sec at 60°C, with fluorescence measured via the SYBR channel after each cycle. Melt curves were assessed post-PCR to ensure no non-specific amplification occurred.

Volumes of 0.2mL (nasopharyngeal swabs) or up to 1mL (FA EBC) of clinical samples collected in Greece were RNA extracted using the Complex800_V6_DSP protocol with the QIAsymphony DSP/Pathogen Midi kit (SafeBlood BioAnalytica SA, Athens, Greece); final elution volumes were 0.06mL. SARS-CoV-2 RNA load was quantified using the VIASURE SARS-CoV-2 (ORF1ab and N genes) Real Time PCR Detection kit for the specific identification and differentiation of SARS-CoV-2 with a manufacturer’s stated analytical limit of detection of 10 target copies per reaction (5μl RNA extract in a 20μl reaction; CerTest Biotec, Zaragoza, Spain), using the internal control at a ratio of 0.1μl internal control/μl eluate. All samples were analysed in single reactions and viral load was expressed as the average Ct of the positive targets per sample.

For maximal sensitivity to SARS-CoV-2 detection in the cohort sampled in Brazil, the whole FA EBC volumes collected were submitted to RNA extraction using PureLink^TM^ RNA Mini Kit, Invitrogen^TM^ (Carlsbad, CA, USA) according to the manufacturer’s instructions; final elution volumes were 30μl. Extract volumes of 2μl were then assayed by real time RT-PCR in 10μl reactions containing iTAQ Universal Probes One-Step Kit, Bio-Rad Laboratories (Hercules, CA, USA) supplemented with pan-eukaryotic 18S rRNA Taqman^®^ primer/probe mix (Thermo Fisher) or the multiplexed CDC SARS-CoV-2 assay (Integrated DNA Technologies Inc., Coralville, IA, USA), targeting two different regions of the viral nucleocapsid gene (N1 and N2 (27)) according to the manufacturer’s protocol. Reactions were carried out in technical triplicates (20% of total sample RNA extract) to increase the chances of detection below the assay limit of quantification (28, 29). Samples were declared positive if all three technical replicates successfully amplified, or inconclusive if at least one of the three technical replicates of each target failed to amplify. Ct’s for each target were calculated as the average of three technical replicates (± standard deviation) with SARS-CoV-2 Ct’s defined as the average Ct’s of the two targets.

### Microbial culture

Blood agar (0.5% peptone, 0.3% yeast extract, 1.5% agar, 0.5% NaCl (Formedium Ltd., Norfolk, UK), 5% defibrillated horse blood (Scientific Laboratory Supplies, Nottingham, UK), pH 7.4) and nutrient agar (0.5% peptone, 0.3% yeast extract, 1.5% agar, 0.5% NaCl, pH 7.4; Formedium Ltd.) plates were poured aseptically and stored at 4°C <1 week. Frozen samples were thawed at room temperature and 0.5mL of sample was pipetted onto the agar plates aseptically and spread using a sterile loop. Plates were incubated at 37°C for 48 hours, after which colonies grown on the plates were photographically documented.

### 16S rRNA microbiota analysis

DNA extraction was undertaken using a modified protocol the Purelink genomic DNA kit (Thermo Fisher Scientific) as previously described (30). Early prototype experiments were performed on an Ion Torrent PGM (Thermo Fisher Scientific) according to the manufacturer’s instructions and as previously described (30). Briefly, Muyzer 16S rRNA V3 primers were used for library generation by direct, extraction/purification free, 40 cycle amplification of either FA EBC or LD added to primer (Integrated DNA Technologies BVBA, Leuven, Belgium) and Kappa Plant polymerase (Merck, Manchester, UK) mastermixes. Aqueous breath sample volumes were used to bring mastermix concentration to 1x in 0.05mL reactions, and samples were barcoded prior library construction and sequencing. Data analysis was performed with QIIME using RDP v11.2 as a reference database; principal coordinate analysis was undertaken using Bray Curtis distances on normalized datasets.

For kitome-corrected microbiomics in PBM-HALE^TM^-collected samples vs RNAse-free water, the same library preparation procedure was adopted, but V4 16S rRNA primers and an Illumina MiSeq (Illumina UK Ltd., Cambridge, UK) platform was used as previously described with post-library construction barcoding (31) at the NU-OMICS DNA sequencing research facility (Northumbria University). Data analysis was performed with Mothur (v1.44.1) using RDP v11.5 and contaminating OTUs were removed using the frequency model in decontam (32). Statistical analysis was performed in R using the phyloseq package to perform alpha and beta diversity measures.

### Metabolomics

All chemicals used in the EBC process and liquid chromatography/mass spectrometry (LC/MS) profiling were Optima grade or equivalent (Thermo Fisher Scientific). The lyophilised EBC samples were reconstituted in 0.05mL of 95/5 v/v acetonitrile and LC/MS water, sonicated for 15min on ice/water bath and centrifuged at 16,580 x g, 4°C for 15min. Supernatants were filtered via 0.22μm Corning Costar Spin-X cellulose acetate spin filters (Thermo Fisher Scientific) and transferred to 1.5 autosampler vials with 0.2mL microinsert. 0.01mL of each sample were pooled together to create a master pool for AcquireX template data-dependent analysis. Extraction blanks were also included to generate the ion excursion list as part of the AqurieX workflow. MS signals associated to the processing workflow were identified using the extraction blanks and an ion exclusion list were generated as part of the data acquisition workflow. Metabolite profiling was performed on a Thermo Fisher Scientific Vanquish Liquid chromatography chromatographic separation system connected to an IDX High Resolution Mass Spectrometer. The Hydrophilic Liquid Interaction Chromatography (HILIC) mode was applied and chromatographic separation was archived using a Waters Acquity UPLC BEH amide column (2.1 x 150mm with particle size of 1.7μm) operating at 45°C with a flow rate of 0.2mL/min. The LC gradient consisted of a binary buffer system, buffer A (95% Water/5% acetonitrile) and Buffer B (95% acetonitrile/5% water), both buffers containing 10mM ammonium formate. Independent buffers systems were used for positive and negative mode acquisition respectively; for positive mode the pH of buffers were adjusted using 0.1% formic acid and for negative 0.1% ammonia solution. The LC gradients were the same for both polarities, T0 95% B at T0 hold for 1.5min and linearly decreased to 50% B, at 12min held for 4.5mins, and returned to starting conditions and held for further 4.5min (column stabilization). The voltage applied for positive mode and negative mode was 3.5kV and 2.5kV respectively. The injection volume used for positive mode was 8μL and negative mode 12μL. The MS data were acquired using the AcquieX acquisition workflow (data dependent analysis using the following operating parameters: MS1 mass resolution 60K, for MS2 30K. Stepped energy (HCD) 20, 25, 50 was applied with a scan range 100-1000, RF len (%) 35, AGC gain intensity threshold was set to 2e^4^ 25%, and custom injection mode was used with an injection time of 54ms. An extraction blank was used to create a background exclusion list and a pooled QC were used to create the inclusion list. The voltage applied for positive mode and negative mode was 3.5kV and 2.5kV respectively. The HESI condition for 0.2mL were as follows: Sheath Gas: 35; Aux Gas 7 and Sweep Gas of 0; Ion Transfer tube Temp: 300°C and Vaporizer Temp 275°C. Post data processing: The HILIC Positive and Negative data sets were processed via Compound Discoverer v.3.2 (Thermo Fisher Scientific) according to the following settings: Untargeted Metabolomic workflow: mass tolerance 10ppm, maximum shift 0.3min, alignment model adaptive curve, minimum intensity 1e6, S/N threshold 3, compound consolidation, mass tolerance 10ppm, RT tolerance 0.3min. Database matching were performed at MS2 level using Thermo Scientific m/z cloud database with a similar index of 70% or better. Only ID metabolite via MS2 were retained.

### Cell culture, virology, and fluorescent cell sorting

HEK293T cells (max passage 20; doubling time of 24hrs) were seeded at a density of 10,000 cells/well in a 96-well plate with culture media consisting of 10% foetal bovine serum (FBS), 1% penicillin/streptomycin Dulbecco’s Modified Eagle Media (Thermo Fisher Scientific) and cultured for 24 hours at 37°C, 5% CO_2_.

Green fluorescent protein (GFP) encoding, vesicular stomatitis virus (VSV) glycoprotein pseudotyped lentivirus (VSV-GFP), rabies virus (RABV), and D614G Beta SARS-CoV-2 were produced as previously described (33). Infectious virus concentration was determined by titration culture on HEK-293T cells followed by fluorescent cell sorting (fCM; BD FACSCanto^TM^ II, BD Biosciences, Wokingham, UK) for GFP according to Grigorov et al (34). Briefly, 0.05mL volumes of virus were plated onto HEK-293T cultures (20,000 cells) and cells were harvested at 72h post-plating using 0.025% Trypsin/EDTA (Thermo Fisher Scientific) and re-suspended in 0.5% FBS in phosphate buffered saline (PBS) after washing once in PBS. Untransduced cell samples were used to create a gate on a side scatter (SSC-A) vs (FSC-A) plot selecting for viable HEK293T cells. A second gate was then created on a fluorescin vs SSC-A histogram, whereby <0.1% of negative control cells were included and thus denoted as GFP-positive.

### Mechanical Nanoparticle and Viral Particle Nebulisation

Nanoparticle suspensions diluted in cell culture media, physiological saline, or deionized water were nebulized for 5-15 min using a PARI Turboboy SX or PARI Boy Classic (PARI Medical Ltd., Byfleet, UK) devices that generate aerosols with a mass median diameter of 3.5μm wherein 67% of the mass is in particles <5μm. Aerosols were routed directly into PBM-HALE^TM^ using a custom 3D printed polylactic acid adapter.

The VSV-GFP stock (8.26×10^5^IFU/mL) was diluted to 5.23×10^3^IFU/mL using reduced serum culture media (2% FBS) whereupon 0.05 mL addition of this 2x working concentration to an equal volume culture 20,000 HEK-293T cells would result in a MOI of 0.0131 and GFP-positive cell counts detectable above background cell autofluorescence with these fCM settings. Volumes of 15mL were then nebulized for 15min, by arresting nebulization at 10min to centrifugally (4000xg for 5min at 4°C) eliminate frothing of the VSV-GFP sample in the nebulization chamber.

Polystyrene flurosphere beads (100nm diameter 505/515 fluorescence FluoroSpheres) were obtained from ThermoFisher. Liposomes were produced by synthetic lipids (Avanti Polar Lipids, Alabaster, AL, USA) using thin-film hydration and extrusion as previously described (35). Lipid composition was either 99 mol% DOPC (neutral liposomes) or 49 mol% DOPC and 50 mol% DOPS (negative liposomes), in each case including 1 mol% TopFluor-PC to enable detection by fluorescence nanoparticle tracking analysis. Lentiviral VLPs (MLVgagYFP-VLPs) and SARS-CoV-2-VLPs were prepared as previously described (36, 37). Nanoparticle size, zeta potential, and concentration were determined using a ZetaView TWIN (ParticleMetrix GmbH, Inning am Ammersee, Germany), at 480nm with the exception of SARS-CoV-2-VLPs, which are not tagged and were analyzed in scatter mode.

### Aerosolised Virus Infectivity Assay

The aerosolized VSV-GFP suspensions collected by PBM-Hale^TM^ were allowed to thaw on ice and centrifuged at 1,000xg for 1min at 4°C to accrue liquid samples. Half the media in 20,000 cell HEK-293T 0.1mL cultures in 96 well plates was then replaced with the aerosol liquid samples to transduce the cells. Supernatants were replaced with fresh culture media at 24h, and GFP positive cells were documented at 72hrs post transduction by fluorescent microscopy (triplicate brightfield and fluorescin channel pictures, BioRad ZOE) at 20x magnification. Nebulised media lacking VSV-GFP were used as a negative control, whereas an aliquot of the non-nebulised 5.22×10^3^ IFU/mL VSV-GFP suspension working dilution was used as the positive control. The fraction of cells positive for GFP expression was then recorded and data was expressed as a % of the mean number of GFP positive cells for the positive control samples, thus providing a value for infectivity relative to the positive control.

### Statistics

Statistical analyses were carried out in GraphPad Prism v9.3.1 (GraphPad Software, San Diego, CA). Sampling consistency was determined by plotting sample volumes over time or number of breaths sampled, and performing regression analyses after anchoring Y axis intercepts to zero and computing the probability of regression slope difference. Differences in culturable microbe load, particle counts/concentrations, and analyte levels were compared by ANOVA followed by post hoc tests for pairwise comparisons (Tukey multiple comparison tests for low replicate virological and qPCR experiments (n=3-5) with limited variable analysis, Benjamini and Yekutieli false discovery rate correction for limited sample (n=3-8) 2-way ANOVA comparisons of operational taxonomic unit (OTU) relative abundance, and Holm-Šídák multiple comparisons for large replicate (aerosolized particle count and NTA) experiments). Paired sample analyses in FA EBC yield, and SARS-CoV-2 viral load vs 18S rRNA levels were conducted by Wilcoxon matched pairs, signed-rank test, and non-parametric Spearman correlation, respectively.

## Results

### The FA EBC microbiome of healthy volunteers resembles that of surgically resected lung tissue

Early experiments were conducted with an EBC collector assembled using laboratory consumables (Figure 1A). Healthy volunteer sampling indicated linear FA EBC volume capture vs the number of tidal oral exhalations (R^2^ 0.8872-0.9960), consistent between consecutive days (slope difference: p=0.8112, n=3; Figure 1B), with highest yields at −78.5°C condensation (Figure 1C). By contrast, no condensation was observable nor aqueous matter retrievable from devices left open to the environment by removing the tube clamps for up to 30min. Under aerobic conditions nutrient agar plating of the FA fraction demonstrated low biomass but culturable bacteria, whose viability was condensation temperature-dependent (Figure 1D). Subjecting LD and FA samples to endpoint PCR for the 16S rRNA gene consistently demonstrated microbial DNA detection, which was free of background contamination (no kitome) only when amplification was performed without nucleic acid extraction or purification (Figure 1E). This finding reflected parallel experiments indicating that freeze/thawing rendered viral nucleic acids directly accessible to PCR (33).

To better understand the microbial diversity of these two sample types, 16S rRNA amplicons generated directly from samples donated by a healthy volunteer over 4 consecutive days, were subjected to Ion Torrent PGM sequencing. The resulting microbiota indicated a stable proteobacteria-dominated (59.1% (±6.75)) signature in FA EBC that was consistently different to the paired, firmicutes-dominated (48.5% (±8.69)) LD samples (Figure 1F). Genus level analysis further indicated that none of the OTUs with a >1% relative abundance in one sample set was present in the other, with Prevotella, Streptococcus, and Veillonella spp. accounting for the LD dominating genera, whilst Enterococcus and Pseudomonas dominated the FA fraction (Figure 1G). Notably, no Haemophilus was present in the FA samples, despite its presence in the LD fraction. These results suggested that this EBC collector design may favour the isolation of oral LD contaminants from respiratory FA.

### PBM-HALE^TM^ enhances FA EBC capture

Key principles behind the laboratory prototype (Figure 1A) were implemented in PBM-HALE^TM^ (WO2017153755 (A1)) whilst introducing changes for improved safety and ease of use (Figure 2A-E; video 1 online demonstrating operational and mechanical principles). These included enlargement of the conducting tubes to 1cm diameter to eliminate tidal airflow resistance, a tamper-resistant coolant chamber, and a one-way valve preventing cooled air inhalation through the device. However, early attempts failed to reproduce FA sampling linearity (Figure S1A) or yield FA samples free of salivary alpha amylase, indicating substantial sample loss or even condensate-driven blockage of the FA platform (Figure S1B), due to component dimensions, material- and device architecture-dependent heat gradients between components, and thermophoretic condensate formation occluding airflow. These were resolved by parametric architecture optimization to achieve high FA sampling linearity across 2-30min tidal breathing sampling periods (R^2^=0.9987-0.9995, n=5 volunteers; Figure 2F).

To better understand how exhaled breath is condensed in PBM-HALE^TM^ we first measured device surface temperature during tidal breathing to note no quantifiable change in condensing surface temperature when sampling at either at 0°C or −78.5°C. Gas phase water nucleates upon surfaces, dry, and aqueous particles at low temperatures (38). We therefore reasoned that swelling fine aerosols in the PBM-HALE^TM^ condenser would experience more gravitational sedimentation and inertial impaction. Computation flow modelling of a typical 500mL tidal oral exhalation through PBM-HALE^TM^ supported this tenet since the FA sample tube rapidly cooled during the inhalation phase, when flow through the device was paused (Figure 2G; video 2 online). To test this hypothesis, we measured tidal exhaled particle counts per second at the PBM-HALE^TM^ exhaust over 3min, as a function of condensation temperature. These experiments showed a reduction in ultrafine particle counts (<0.3 μm; p<0.01) on account of 0°C or −78.5°C condensation, matched by simultaneous increases in 0.5-10 μm particle sizes (p<0.05 to 0.0001), with a consistent trend supporting a condensation temperature-dependent effect on particle size (Figure 2H-M). Taken together these results indicated that the FA mass was most effectively cooled, allowing FA to swell in size and condense onto PBM-HALE^TM^ during the inspiration phase of the breath cycle.

### FA EBC consistency in healthy volunteers sampled with PBM-HALE^TM^

As with early experiments, FA EBC aerobic microbe viability was condensation temperature-dependent (n=3; Figure 3A); however, continuous sampling could not extend beyond 40 min due to loss of coolant contact with the condensation surface (n=5; Figure S1C). Furthermore, LD microbial load was lower than that of drooled saliva, indicating that exhaled LD may consist of salivary and respiratory droplets. Accordingly, salivary alpha amylase activity was lower in the LD fraction vs drooled saliva (n=5; Figure 3B); yet neither 5 exhalations (Figure S1D) nor 30 mins of continuous sampling yielded any assayable salivary alpha amylase activity in the FA fraction (<0.4units/mL; n=5), even after 10x sample concentration by lyophilisation. Taken together, these data reinforced the premise that PBM-HALE^TM^ inertially separates orally contaminated LD for effective lower respiratory tract FA EBC condensate collection.

**Figure 3:**
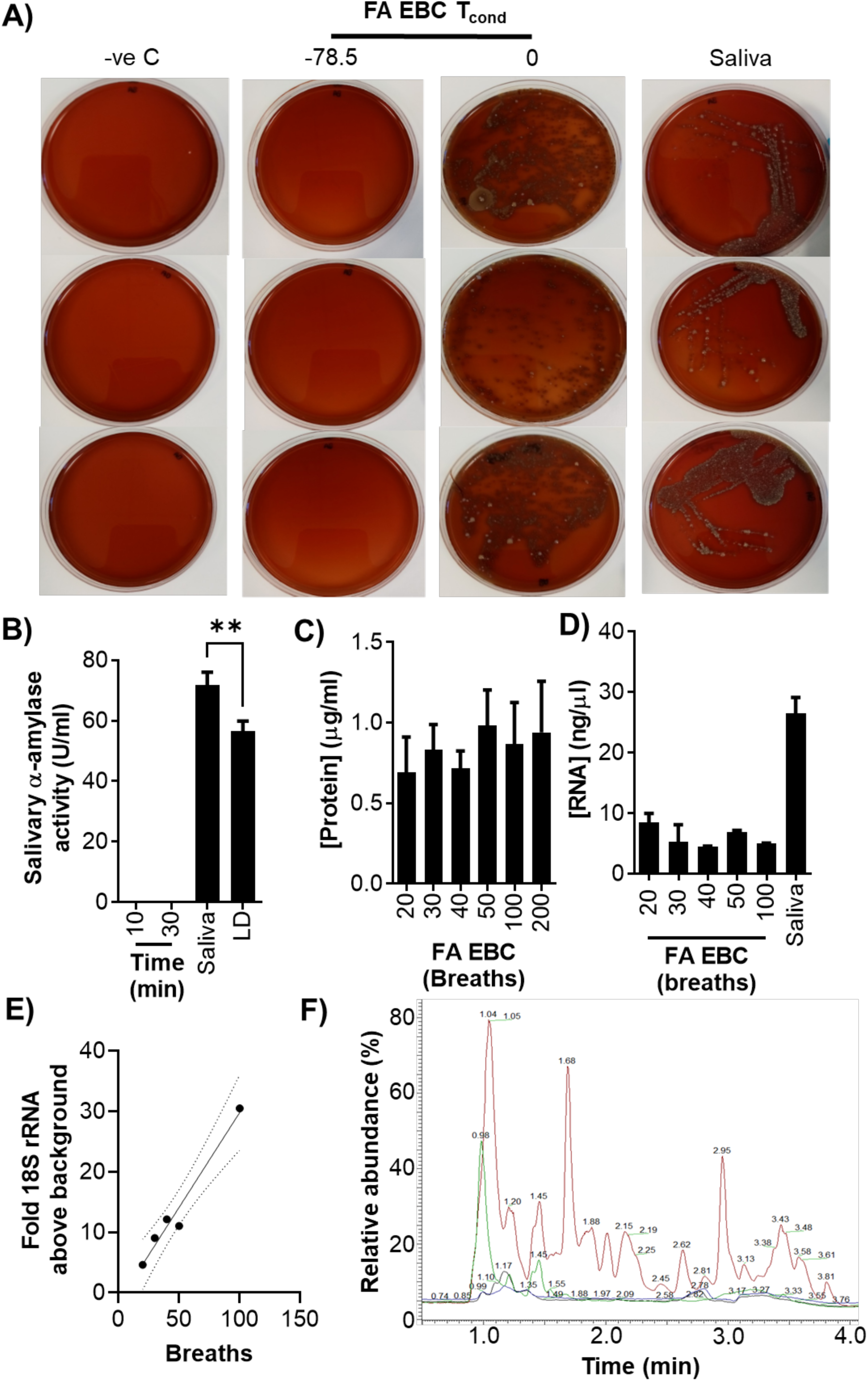
Bacterial and biomolecular composition of FA EBCs. (A) Aerobic bacteria viability in saliva samples and 2min FA EBC samples as a function of condensation temperature (T_cond_), as determined by blood agar plating. (B) Salivary α-amylase levels in saliva, LD fractions, and FA EBC samples captured for 10 and 30min at −78.5°C condensation from a healthy male adult volunteer (n=5). C) Protein concentration in FA EBC samples captured for an increasing number of breaths as determined after RIPA buffer extraction and microBCA assay (n=5). D) Spectrophotometric determination of RNA concentration in extracted FA EBC samples captured for an increasing number of breaths from an adult healthy male volunteer (n=5). E) Eukaryotic 18S rRNA levels in extracted RNA from FA EBC samples captured from an adult male healthy volunteer for 20-100 breaths (duplicate technical replicates plotted). F) Representative HILIC-MS plot of metabolites detected in 10min FA EBC samples from an adult male healthy volunteer. blue: direct injection negative control; green: acid extraction negative control; black: direct injection of FA EBC; red acid-extracted FA EBC. Histograms in B, C, and D show averages ± standard deviation.

Focusing therefore analyses on the tidal FA fraction, healthy volunteer samples featured DNA quantities at, or below the limit of detection, and low cell-free protein content, both quantifiable only after 5x lyophilization concentration (∼0.833 (±0.117)μg/mL; n=5). In contrast, there were appreciable RNA quantities (6.03 (±1.61)μg/mL; n=5), at concentrations stable irrespective of sampling duration (Figure 3C,D). RT-qPCR analysis indicated 5.21x (±0.88) above-background enrichment (p<0.001; n=5; Figure S1E) for eukaryotic 18S rRNA in 20 tidal breath samples, rising to 9,432x (±923) for a 30 min sample, with total yield correlating to sampling period length (R^2^=0.9677; Figure 3E). In contrast, human GAPDH or beta actin mRNA were below assay detection limits. Interestingly, 260/280nm RNA absorbance ratios (1.52 (±0.09); n=5) indicated the presence of substantial volatile contaminants co-extracted with FA EBC RNA, as parallel extraction of equal volumes of saliva presented solvent-free RNA (A_260/280_=1.8 (±0.012); n=3).

Given the established volatile compound content in exhaled breath, we next examined organic analyte content by HILIC-MS. Unlike PCR, direct injection of a 10min, 0.5mL sample resulted in no appreciable peaks; however, injection of extracted samples (n=5) yielded 141 non-volatile compounds <837.5g/mol (phosphatidylethanolamine), of which 20 resulted in multiple hits in the human metabolome database, and 104 were novel compounds (Figure 3F; Table 1). The most prevalent unique hits included n-decanoylglycine and n-nonanoylglycine among several other eukaryotic cell membrane lipids whose lysis-dependent detection alluded to the presence of intact cells or extracellular vesicles. Other interesting compounds included N-acetylputrescine, a urea cycle byproduct commonly causal of halitosis whose serum elevation is implicated in lung cancer (39) and the pyrrolidine derivative cuscohygrine. This is a coca leaf alkaloid previously presumed to be lost during cocaine fabrication (40) and detected in oral fluids of coca leaf chewers and coca tea drinkers but not cocaine users (41). In this instance cuscohygrine provenance was on account of cocaine nasal consumption ∼72hrs ahead of EBC sample collection with PBM-HALE^TM^indicating that cocaine abuse might be detectable in FA EBC (40).

**Table 1:** List of annotatable metabolites detected in extracted FA EBC from an adult healthy male volunteer after 10min of sampling.

| **Compound** | **RMM\***<br>**(g/mol)** | **RT<sup>†</sup>**<br>**(min)** | **Relative ion**<br>**abundance** |
| --- | --- | --- | --- |
| 1-alkyl-sn-glycerol 3-phosphate | 396.3 | 1.002 | 810,094 |
| 1-Acylglycerol | 352.3 | 1.020 | 281,866 |
| Lysophosphatidic acid | 410.2 | 1.032 | 968,316 |
| Palmitoleoyl ethanolamide | 297.3 | 1.047 | 187,282 |
| Arachidonic acid | 335.2 | 1.054 | 348,544 |
| Linoleamide | 279.3 | 1.061 | 216,809 |
| Cuscohygrine | 224.2 | 1.067 | 723,759 |
| N-decanoylglycine | 229.2 | 1.156 | 2,612,124 |
| N-Nonanoylglycine | 215.2 | 1.198 | 1,942,872 |
| cis-3-Hexenyl b-primeveroside | 394.2 | 1.221 | 160,089 |
| N-lauroylglycine | 257.2 | 1.923 | 286,977 |
| N-undecanoylglycine | 243.2 | 2.072 | 227,826 |
| 1,2-distearoylphosphatidylethanolamine | 837.5 | 2.388 | 381,518 |
| Gambogic acid | 628.3 | 2.536 | 416,778 |
| 2-hexenoylcarnitine | 257.2 | 3.062 | 994,821 |
| L-arginine | 175.1 | 3.367 | 502,141 |
| N-acetylputrescine | 130.1 | 3.519 | 192,382 |

\* Relative molecular mass
† Retention time

### PBM-HALE^TM^ captures viruses and nanoparticles suspended in fine aerosols

With the emergence of SARS-CoV-2 we promptly sought to determine if PBM-HALE^TM^ could capture aerosolized viruses in the FA EBC fraction. We first established a BSL2 model of aerosolized virus capture based on GFP-expressing, VSV glycoportein-pseudotyped lentivirus, a similarly sized (∼150nm), enveloped RNA virus (Figure S2AB). Exposing HEK-293T cells to nebulized VSV-GFP passively routed via PBM-HALE^TM^ indicated virus infectivity was reduced as a function of condensation temperature (Figure 4AB). Thus, in line with previous reports with MS2 coliphage and Ebola virus (33) and unlike exhaled aerobic bacteria (Figures 1A, 3A), exhaled viruses may not be effectively disrupted by a single freezing/thawing cycle to support direct detection by RT-PCR, or the handling of FA EBC samples as non-infectious.

**Figure 4:**
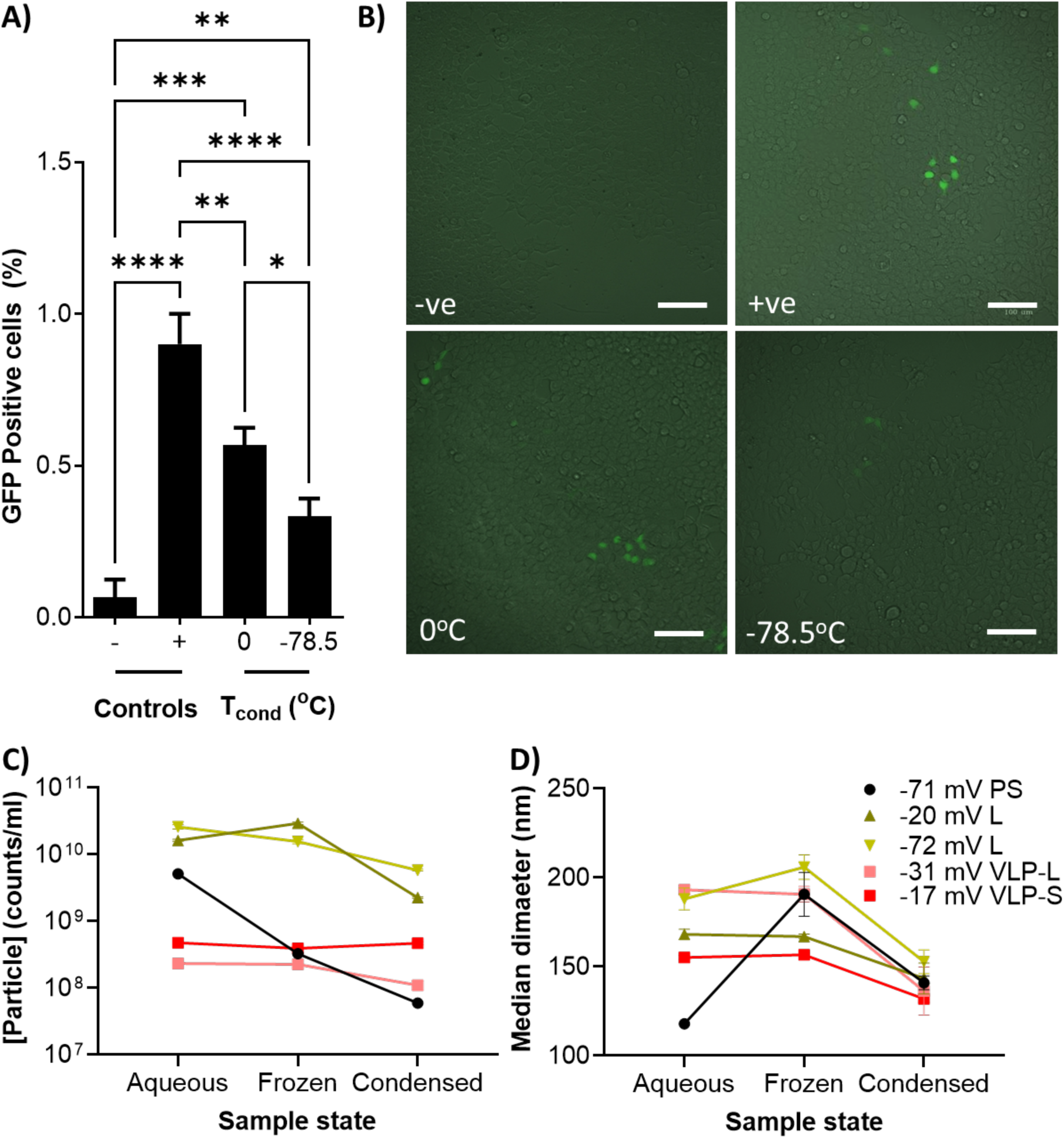
Airborne nanoparticles, liposomes, virus-like particles, and infectious viruses can be captured in the FA fraction of exhaled breath. (A, B) GFP-expressing HEK293T cell detection by fluorescent cell sorting (A) or fluorescence microscopy (B) after mechanical nebulisation of VSV-G pseudotyped GFP-encoding lentivirus and aerosol capture by 0°C or −78.5°C condensation using PBM-HALE^TM^, as compared to direct cell transduction with the nebulization suspension (positive control; +ve) or autofluorescence (negative control; −ve). (C, D) Effect of flash-freezing or mechanical aerosolization/condensation capture of polysterene beads (PS), liposomes (L), lentiviral virus-like particles (VLP-L), or SARS-CoV-2 virus-like particles (VLP-S) on (C) particle concentration and (D) median diameter, as measured by nanoparticle tracking analysis (average ± standard deviations of 3 independent replicates). The net zeta potential (charge) of each particle type is shown. *: p<0.05; **: p<0.01; ***: p<0.001; ****: p<0.0001.

To better understand how nebulization, condensation, and particle charge interact in affecting nanoparticle structural integrity, we repeated these experiments using synthetic polystyrene beads (PS; −71mV, 118nm), negatively charged liposomes (−72 mV, 188nm), neutral liposomes (−20 mV, 168nm), lentiviral virus like particles (VLP; − 31 mV, 193nm) and SARS-CoV-2 VLPs (−17mV, 155nm), measuring the resulting changes in particle size and concentration (Figure 4CD, Figure S2C). In line with the VSV-GFP infectivity assay, lentivirus VLP concentration was reduced by 43.7% with a corresponding reduction in particle diameter of 29.4% on account of nebulization (p<0.0001), but not by freezing of the suspension (2.8% reduced concentration, 1.3% reduction in mean size, n=3). Interestingly, no appreciable change in SARS-CoV-2 VLP particle concentration (p>0.999) was observed indicating that PBM-HALE^TM^ capture of SARS-CoV-2 would have minimal impact on virion integrity. It must be noted that SARS-CoV-2-VLPs, unlike lentiviral VLPs, were not fluorescently tagged and tracked by scatter-NTA instead of F-NTA and non-VLP particles may thus be included in this analysis. By stark contrast, negatively charged nanoparticle concentration in PBM-HALE-captured condensates was reduced by 72.6x-98.0x compared to source fluids (p<0.0001), likely on account of freezing-driven aggregation, in a nanoparticle type-dependent manner (PS beads (p<0.0001) > liposomes (p=0.0031)). Taken together, these results supported the evaluation of tidal breath SARS-CoV-2 airborne transmission using PBM-HALE^TM^.

### Respiratory pathogen detection in tidal oral FA EBCs of symptomatic patients

The distal lung origin of bulk exhaled FA mass (3) motivated us early in the pandemic to ask whether tidal exhaled breath might be a source of airborne SARS-CoV-2. We therefore sampled COVID-19 patient tidal oral exhalations using PBM-HALE^TM^ for up to 30 min and evaluated FA EBC fractions for SARS-CoV-2 by RT-PCR. The first 12 cases involved hospitalized patients either convalescent and nasopharyngeally negative for SARS-CoV-2 (n=2; 3^rd^ week from symptom onset), or within their first 5 days of hospitalization (n=10; >2 weeks after symptom onset). All were sampled for 30 min in COVID-19 wards across 2 hospitals with no HEPA filtration or mechanical ventilation, settings known to be heavily contaminated with airborne SARS-CoV-2 nucleic acid (42). None returned any evidence of SARS-CoV-2 genomes in FA EBC, as the only a single sample with an inconclusive readout (Ct 38 in one of two targets only) was negative on repeat testing. Given cotemporally accruing evidence of nasopharyngeal viral load and transmission declining within the first 5 days of symptom onset (43), we next recruited 30 acutely symptomatic patients (Table 2) confirmed nasopharyngeally positive for SARS-CoV-2. No SARS-CoV-2 nucleic acid was detected among these 1.18mL ± 0.32 FA EBC samples either, despite nasopharyngeal loads as low as Ct 13.1.

**Table 2:** Participant characteristics in the tidal FA EBC SARS-CoV-2 viral load study.

| Variable |  | Overall |  |
| --- | --- | --- | --- |
| Number of participants (N) |  | 30 |  |
| Age (years, ±SD) | | 47.2 | 17.1 |
| Age groups, years (N, %) |  |  |  |
|  | <18 | 0 | 0.00% |
|  | 18-69 | 23 | 76.7% |
|  | 70-80 | 7 | 23.3% |
| Sex (N, %) |  |  |  |
|  | Male | 15 | 50.0% |
|  | Female | 15 | 50.0% |
| Height (cm, ±SD) | | 171 | 6.75 |
| Weight (kg, ±SD) | | 73.5 | 21.8 |
| BMI |  | 25.2 | 8.12 |
|  | <18.5 | 2 | 6.67% |
|  | 18.5-24 | 14 | 40.0% |
| | ≥24 | 16 | 45.7% |
| Armspan (cm, ±SD) | | 172 | 7.82 |
| Nasopharyngeal viral load |  |  |  |
| (Ct, ±SD) | | 21.2 | 5.43 |
| Days after symptom onset |  |  |  |
| (N, ±SD) | | 3.17 | 1.09 |
| Symptoms (N, %) |  |  |  |
|  | Fatigue | 20 | 66.7% |
|  | Dyspnoea | 5 | 16.7% |
|  | Diarrhoea | 2 | 6.67% |
|  | Cough | 10 | 33.3% |
|  | Rhinorrhoea | 1 | 3.33% |
|  | Headache | 7 | 23.3% |
|  | Muscle pain | 11 | 36.7% |
|  | Joint pain | 1 | 3.33% |
|  | Dysgeusia/Ageusia | 4 | 13.3% |
|  | Dysosmia/Anosmia | 6 | 20.0% |
| Fever (N, %) |  | 22 | 73.3% |
|  | Mild (<38oC; N, %) | 17 | 77.3% |
|  | Moderate (38oC-40; N, %) | 5 | 22.7% |
|  | Severe (>40oC; N, %) | 0 | 0.00% |
| Other lung disease |  |  |  |
|  | Asthma (N, %) | 1 | 3.33% |
|  | Chronic bronchitis (N, %) | 0 | 0.00% |
|  | Emphysema (N, %) | 1 | 3.33% |
|  | COPD (N, %) | 1 | 3.33% |
|  | Tuberculosis (N, %) | 0 | 0.00% |
|  | Lung cancer (N, %) | 0 | 0.00% |
|  | Lung fibrosis (N, %) | 1 | 3.33% |
| Comorbidities (N, %) |  | 7 | 23.3% |
| Lung Surgery (N, %) |  | 0 | 0.00% |
| Inpatient (N, %) |  | 19 | 63.3% |

To determine whether bacterial causes of symptomatic respiratory infections could be detected by FA EBC, we analysed the tidal oral FA EBC microbiome of an acutely symptomatic male with a mild dry cough, negative by nasopharyngeal RT-PCR for SARS-CoV-2. Illumina V4 16S rRNA profiling by direct amplification of 10-50 exhalation samples indicated above background bacterial DNA enrichment and only four OTUs as the genera overrepresented across all FA EBC samples after FDR correction (Figure 5AB). Of these, only Streptococcus and Corynebacterium spp. were entirely absent from negative control samples, and only Streptococcus spp. were found at relative frequencies consistently significantly different to negative controls irrespective of sampling duration (q<0.0001 to 0.0164), and therefore the most likely cause of symptoms (Figure 5C).

**Figure 5:**
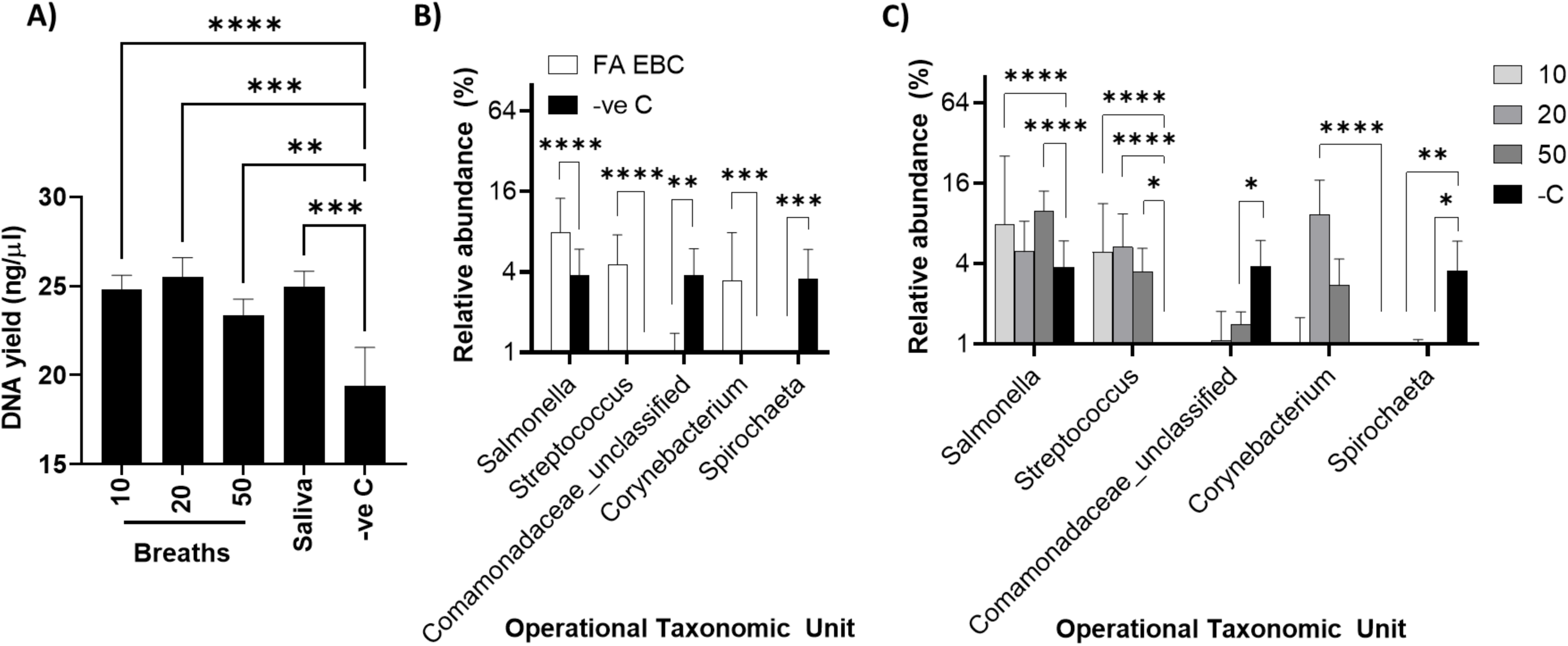
Detection of *Streptococcus* in the FA EBC of a case with a mild cough by direct 16S amplicon generation from FA EBC and next generation sequencing. (A) Quantification of 16S rRNA V4 amplicon library yields from FA EBCs collected as a function of an increasing number of breaths (n=5), and saliva (SAL; n=3), vs negative control (-ve C; RNase free water; n=5) after direct sample amplification with Kappa Plant 3G polymerase without extraction/purification (means ± standard deviation). (B, C) Comparison of genus-level OTU relative abundance across all FA EBC samples vs all negative control samples (B) or by duration of breath sampling (C) identifies *Streptococcus, Corynebacterium and Salmonella* spp. enrichment in the patient FA EBC, with only *Streptococcus* spp. being significantly enriched across all timepoints (n=5). Data show average ± standard deviations *: p or q<0.05; **: p or q<0.01; ***: p or q<0.001; ****: p or q<0.0001.

To evaluate whether our sampling process might disfavor SARS-CoV-2 capture, or compromise virion/genome stability, we assessed the effect of condensation temperature on sample volume and concentration. Thus, a 13.9x increase in 30 min FA EBC volumes on account of −78.5°C condensation was observed (Figure 6A), indicative of condensation-driven dilution. The resulting osmotic shock upon sample thawing could therefore expose viral genomes to RNase activity explaining failure to detect SARS-CoV-2 RNA in COVID-19 patients. However, 14x VSV-GFP dilution in 18 MOhm water, flash freezing, and room temperature incubation for up to 24hrs resulted in no appreciable impact on virus infectivity (Figure 5B-D). At the same time, −78.5°C condensation resulted in only a 2.2x drop in FA EBC extracted protein concentration (Figure 6E), suggesting an enhancement of FA EBC capture by more than 6x on account of particle swelling, rather than simply sample dilution or microbial cell, viral particle, or vesicle lysis. Taken together our in vitro, healthy volunteer, and clinical observations indicate that absence of detectable SARS-CoV-2 in tidal FA EBC of acutely symptomatic patients is not an artefact of the sampling or analytical process but a function of viral load, if any, being <120 genome copies per mL of FA condensate (4.72 genomes/min).

**Figure 6:**
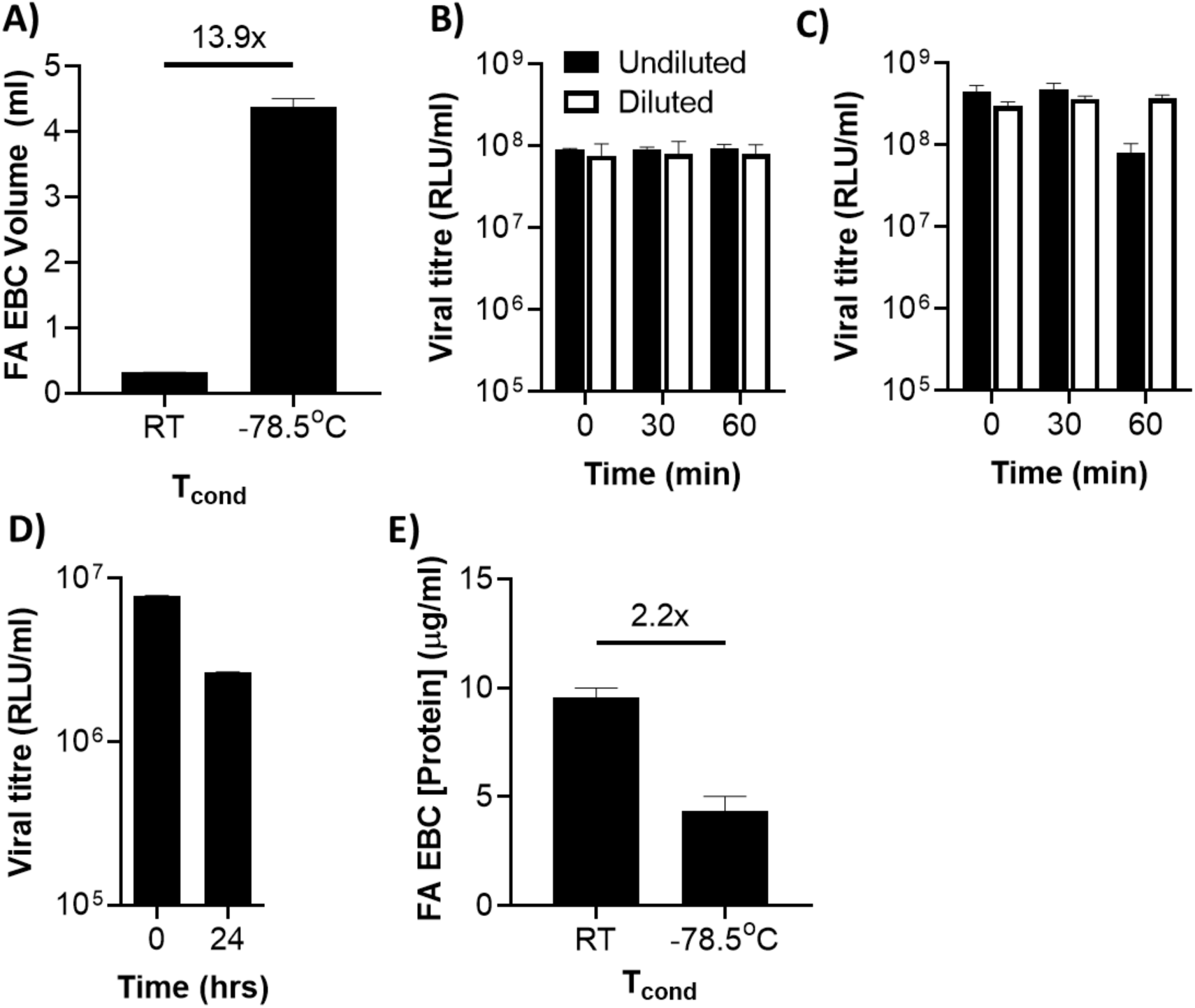
Vapour condensation-driven FA swelling and dilution enhances protein capture without affecting viral infectivity. (A) FA EBC volume as a function of condensation temperature after sampling a healthy adult female volunteer for 30min (n=3, average ± standard deviation). (B, C) Two pseudotyped lentiviruses, a GFP-reporter D614G SARS-CoV-2 spike-pseudotyped lentivirus (B) or rabies glycoprotein-pseudotyped lentivirus (C) were incubated for 0, 30, or 60min after 1:14 dilution in PBS or 18 megaOhm water, and were subsequently seeded onto HEK293T cells for 72 hours to assess impact on infectivity. (D) GFP-reporter Beta variant SARS-CoV-2 spike-pseudotyped lentivirus was incubated at room temperature for 24hrs and impact on HEK293T cell infectivity was compared to 4°C storage. Virus titres were calculated as the mean relative light units/min (n=3, average ± standard deviation; data representative of 3 independent experiments). (E) Total extracted protein content in FA EBC as a function of condensation temperature after sampling a healthy adult female volunteer for 30 min (n=3, average ± standard deviation).

### Forced expiration enables SARS-CoV-2 detection in FA EBCs of symptomatic patients

Recent reports (44, 45) suggested SARS-CoV-2 load may increase in exhalations as a function of vocalization intensity. These data motivated us to evaluate changes in FA EBC viral load in a small cohort of acute cases (Table 3) as a function of tidal vs forced expiration. Patients were recruited among attendees of a suburban primary care centre, and sampling was conducted in a highly naturally ventilated screening room. Quantification of total FA EBC mass indicated that FA EBC yield rates did not differ by exhalation mode (0.121mL ±0.0471 vs 0.106mL ±0.0462; p = 0.6387; Figure 7A). This was expected considering that only the terminal 48mL of each breath is effectively condensed by PBM-HALE^TM^ (Figure 2G) and not the entire sample. However, only forced expiration yielded normalized distribution data, indicating additional factors may influence tidal FA generation in the distal lung between individuals. Accordingly, no correlation was observed between tidal and forced expiration FA production rates (p=0.7925).

**Table 3:** Participant characteristics in the tidal vs forced expiration FA EBC SARSCoV-2 viral load study.

| Variable |  | Overall |  |
| --- | --- | --- | --- |
| Number of participants (N) |  | 15 |  |
| Age (years, ±SD) | | 47.7 | 11.4 |
| Age groups, years (N, %) |  |  |  |
|  | <18 | 0 | 0.00% |
|  | 18-69 | 15 | 100% |
|  | 70-80 | 0 | 100% |
| Sex (N, %) |  |  |  |
|  | Male | 4 | 26.7% |
|  | Female | 11 | 73.3% |
| Height (cm, ±SD) | | 166 | 6.91 |
| Weight (kg, ±SD) | | 76.8 | 18.7 |
| BMI |  | 27.9 | 5.80 |
|  | <18.5 | 0 | 0.00% |
|  | 18.5-24 | 4 | 26.7% |
| | ≥24 | 11 | 73.3% |
| Armspan (cm, ±SD) | | 161 | 13.9 |
| Nasopharyngeal viral load |  |  |  |
| (Ct, ±SD) | | 22.5 | 2.75 |
| Days after symptom onset |  |  |  |
| (N, ±SD) | | 3.53 | 1.06 |
| Symptoms (N, %) |  |  |  |
|  | Fatigue | 5 | 33.3% |
|  | Dyspnoea | 0 | 0.00% |
|  | Diarrhoea | 2 | 13.3% |
|  | Cough | 11 | 73.3% |
|  | Rhinorrhoea | 15 | 100% |
|  | Headache | 11 | 73.3% |
|  | Muscle pain | 7 | 46.7% |
|  | Joint pain | 2 | 13.3% |
|  | Dysgeusia/Ageusia | 2 | 13.3% |
|  | Dysosmia/Anosmia | 2 | 13.3% |
| Fever (N, %) |  | 0 | 0.00% |
|  | Mild (<38oC; N, %) | 0 | 0.00% |
|  | Moderate (38oC-40; N, %) | 0 | 0.00% |
|  | Severe (>40oC; N, %) | 0 | 0.00% |
| Other lung disease |  |  |  |
|  | Asthma (N, %) | 1 | 6.67% |
|  | Chronic bronchitis (N, %) | 1 | 6.67% |
|  | Emphysema (N, %) | 1 | 6.67% |
|  | COPD (N, %) | 1 | 6.67% |
|  | Tuberculosis (N, %) | 0 | 0.00% |
|  | Lung cancer (N, %) | 0 | 0.00% |
|  | Lung fibrosis (N, %) | 1 | 3.33% |
| Comorbidities (N, %) |  | 2 | 13.3% |
| Lung Surgery (N, %) |  | 0 | 0.00% |

**Figure 7:**
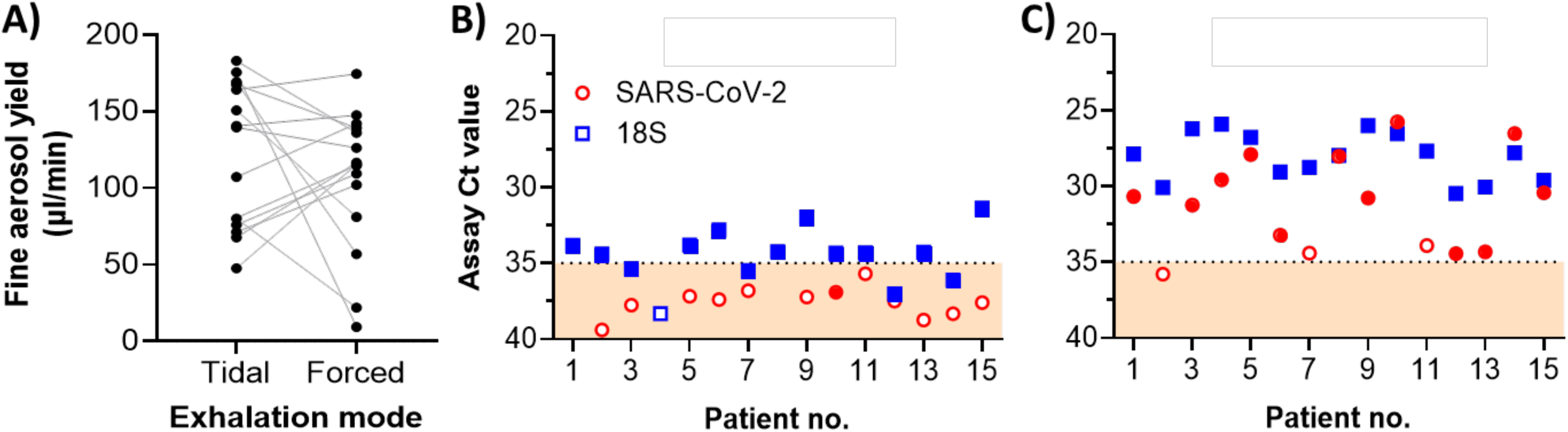
Forced expiration allows up to 100% detection and quantification of SARS-CoV-2 RNA in EBC of distal lung fine aerosols. (A) Paired sample analysis of FA EBC volume yield per min after 30 min tidal or 15 min forced exhalation (n=15). (B, C) Total RNA content and SARS-CoV-2 RNA levels in acute COVID19 patient FA EBC collected during (B) 30 min tidal expiration or (C) 15 min forced expiration, as measured by 18s rRNA and multiplexed CDC real time RT-PCR. Each sample was analysed in technical triplicates and the resulting average Ct ± standard deviation is shown (Filled datapoints: successful amplification in 3/3 technical replicates and positive for SARS-CoV-2; hollow datapoints: no amplification in at least 1/3 of technical replicates in either of the N1 or N2 assay and inconclusive for SARS-CoV-2; Dotted line and beige area: amplification region below the CDC SARS-CoV-2 assay limit of quantification (Ct = 35); all plate positive and negative controls performed as expected).

We next analysed total RNA content and SARS-CoV-2 genome presence by 18S rRNA and CDC assay real time RT-PCR (27), respectively. The results corroborated previous findings (Figure 3E) regarding 18S rRNA levels in tidal FA EBC (Ct 34.5 ±1.80, i.e. 5.5 cycles above background). Forced expiration, however, increased 18S rRNA concentration by 84.5x (Ct 28.1 ±1.57) indicating sample enrichment with distal airway contents. In support of this tenet, forced expiration yielded 12/15 (80%) SARS-CoV-2 positive samples with a Ct as low as 25.8; the remaining 3 were classed as inconclusive but were detectable at, or near the assay limit of quantification (Figure 7); in the context of symptomatic disease, these results would classify these three patients as likely SARS-CoV-2 positive. By stark contrast, 3/15 tidal breath samples were negative for SARS-CoV-2, whereas the single positive and 11 inconclusive samples were all detected below the assay limit of quantification. Interestingly, 18S Ct values correlated to SARS-CoV-2 Ct values in FA EBC collected by forced expiration (p=0.0213), but not by tidal breathing (p=0.700). These findings suggested forced expiration enriched distal airway content in the resulting FA EBC, enabling non-invasive SARS-CoV-2 RNA quantification in this locus.

## Discussion

### Sampler architecture impacts EBC consistency

The key problems of sampling inconsistency, salivary contamination, and sample loss among EBC collectors (3, 14) gave rise to castigating criticisms (46) over EBC biomarker associations with disease; these persist, restricting clinical translation of this methodology (47, 48). Many of these problems relate to the architecture of EBC collectors. For example, unprotected sampling surfaces such as petri dishes, filter modules, or indeed open-ended condensing tubes risk ambient aerosol condensation and manual handling contamination. Some instruments additionally fail to capture any EBC from some subjects: as in critical care ventilation systems, elephant trunk connectors result in aerosol ‘rain out’ before substantial aerosol quantities reach the EBC sampling vessel. Nevertheless, reports of high natural variability in exhaled aerosol particle load (49) and relative humidity (50) could also explain such observations. Conversely, some solutions rarely integrate any physical means of salivary droplet separation, resulting in substantial salivary contamination (51, 52). Importantly, this risk can be missed if poorly sensitive analytical methods are used (53). Yet other devices, such as the BioScreen I (54) and II (55), can easily suffer saliva droplet contamination in conducting tubing. ‘Fluid film burst’ (1) disruption of these droplets may generate contaminating salivary FA, and in some cases the large droplets can be even propelled into the condenser, especially if tubing is angled downwards (56, 57).

We sought to address EBC sampling inconsistency, salivary contamination, and sample loss by a) preventing sample loss in non-condensing instrument parts; b) physically isolating the condensing surface to prevent environmental contamination or evaporative sample loss; and c) implementing basic principles from inhaled therapeutics design (58) to separate saliva-laden LD from the FA. This approach demonstrated good FA EBC volume linearity vs. duration of use, further improved by eliminating flow resistance. It is noteworthy that small changes in device architecture proved critical, underscoring how EBC collector design impacts sample integrity. Similar observations have been reported by others who have successfully reduced salivary contamination by diverting exhaled breath flow (52). In the case of PBM-HALE^TM^ however, any contaminating saliva in the FA fraction was below salivary alpha amylase assay detection limits, or > 1,750x diluted by FA.

Dilution is a feature of condensation due to the nucleation of exhaled water vapour on condensers, leading to recommendations of 500x sample concentration to detect analytes reliably (51). However, in PBM-HALE^TM^ the resulting dilution is only ∼14x since it drives exhaled particle enlargement and capture increase by ∼6x. This phenomenon is likely universal among EBC condensers where breath remains static in the condenser during the inhalation phase, however condensation temperature decay in some systems will reduce exhaled particle swelling and thermophoresis as condenser temperature rises. Whilst dry ice cooling can sustain condensation temperatures at −78.5°C unperturbed for 30 min of continuous use, the optimal condensation temperature in active cooling solutions remains to be determined and should factor energy requirements vs fine aerosol capture efficiency.

### Microbiome and metabolome analysis in EBC

Exhaled breath and invasive lung sample microbiomics remains a technically challenging, non-standardized field. The only comprehensive study on EBC microbiomes to date did not find stable communities among technical replicates of EBC collected with R-Tube devices that were subjected to DNA extraction (59). In that study, community diversity ‘noise’ and inconsistency was common to negative control DNA extractions, unless >2000 16S rRNA gene copies were present per sample- a near-universal background (kitome) bacterial DNA contamination level (59). Past experience with oral microbiome contamination in very low biomass-containing liquid animal samples with viable aerobes (30) indicated that salivary OTUs would be readily detectable in a contaminated FA fraction. We had also found that at least 8-10 freeze-thaw cycles were necessary for direct amplification of viral RNA in reactions templated with fresh blood (33), yet aerobic bacterial viability in FA EBC was lost in a single freeze-thaw cycle. This motivated us to explore direct 16S rRNA amplicon sequencing library preparation to avoid extraction kit contamination observed by us (Figure 1E) and others (60). Prevotella, Veillonella, and Streptococcus spp. in LDs with this reduced kitome contamination method were consistent with firmicutes and bacteroides (61) results from invasively obtained lung samples, which are subject to the persistent risk of oropharyngeal contamination even in protected brush specimens (62). A ∼60% proteobacteria-dominated FA EBC signature, by contrast, has only been reported in surgically obtained lung resections (63). By taking an extraction-free approach, therefore, kitome contamination is restricted to the sequencing library polymerase, water source, and operator technique. However, in line with the recommendation of Erb-Downward et al., we find technical replicates facilitated above-background respiratory bacterial pathogen detection reliability. Further work is necessary, including an assessment of how exhalation maneuvers affect breath aerosol production and consistency, the impact of infection location, mucous production, mucosal surface hydration, and dyspnoea (64–68).

Unlike reports with devices prone to salivary contamination employing comparable analytical methodologies, metabolite and protein content in FA EBC was restricted to at least vesicular material. Whether the membrane compounds detected are of human or microbial origin is still unclear, but past reports of exosomes in EBC (69, 70) and our observation of lung surfactant components point towards the host as their source. Indeed, exosomes often lack housekeeping gene-coding RNA but are rich in rRNA (71) in line with our observations; future metatranscriptomic studies will establish the diversity and origin of FA EBC RNA and whether this includes miRNAs which are established biomarkers for cancer (72–75), lung inflammation (76–78), and a number of autoimmune disorders (79–81).

### Pinpointing the source of airborne SARS-CoV-2 in exhaled breath

The debate on SARS-CoV-2 airborne transmission (4, 82, 83), often the only explanation for superspreader events (84–87), and the ∼30% false negative error rate of nasopharyngeal swab RT-PCR (88, 89), have motivated several studies on breath-mediated airborne transmission. Their outcomes vary from 0% (56) to 93% (90) of cases nasopharyngeally positive for SARS-CoV-2 yielding breath samples also positive for the virus. Of those returning positive data several involve devices previously reported (90) or readily prone (54–56, 91) to salivary contamination, an established source of SARS-CoV-2 (92), with the important exceptions of the work of Feng et al. and Huang et al. In the first study, a 35L breath and air mixing container was connected on its upper surface to a pump, routing FA into a NIOSH bioaerosol collector, thereby inertially excluding LDs. In the context of this diluting collection method, FAs produced by tidal breathing were reportedly devoid of SARS-CoV-2 genomes. However, ’classical’ EBC collected using a 15mL tube with a cut off tip conducting to a 50 mL tube placed below the 15mL tube, and immersed in coolant, lacking any environmental or salivary contamination prevention, was positive in 25% of cases. In the second study, negative breath samples were also matched by salivary negative samples in patients that were positive by nasopharyngeal swab pointing to saliva as the source of ‘breath’ SARS-CoV-2. Elsewhere, electret filter-based testing did return high concordance in exhaled breath viral load vs nasopharyngeal swabs (93). Yet this technology does not prevent large droplet contamination of the breath specimen filter and has only been explicitly tested for methadone capture efficiency, as opposed to particle size filtering (93, 94).

A salivary source cannot be excluded either from the important reports of Coleman et al. (45) and Adenaiye et al. (44) that established airborne transmission, wherein infectious SARS-CoV-2-laden exhalations were detected in tidal breath, rising in concentration by vocalization intensity. These studies relied on pumping patient exhalations at a 130L/min fixed rate through a ∼1m long cone surrounding the patient’s head, and into an aerosol capture array (95). The system relies on humidified air supply at the cone perimeters which in Coleman et al. was ambient, 68% relative humidity HEPA-filtered air in a COVID-19 ward (Coleman K, personal communication). This reduced the likelihood of ambient contamination, but not necessarily fomite contamination from the subject’s hair or skin. Moreover, the device did not dynamically adjust sampling rates in response to patient exhalation rates e.g. during singing vs tidal breathing. This could result in higher inflow from the cone perimeter, potentially increasing fomite and ambient contamination risk. Perhaps more importantly the cone path length allows for aerosol particle size distribution to evolve in response to the sharp temperature and humidity gradients from the respiratory tract to atmosphere. Ambient air influx and mixing within the cone would exacerbate this effect (96). In their seminal paper on this sampling method, McDevitt et al (95) reported no ambient air FA particle size changes on account of the device cone and pump operation, but <50% capture efficiency for mechanical aerosol particles <30μm; no particle data was reported for human breath aerosols which differ in humidity and temperature to environmental aerosols.

Our study is limited by the lack of evidence of viral loads in the salivary amylase-containing LD fractions and the absence of infectivity evidence for the FA EBC samples obtained. However, by focusing on the distal airway fraction we have confirmed that aerosols produced in the lung during tidal breathing contain negligible quantities of SARS-CoV-2. For tidal breathing alone to yield virus-laden airborne aerosols at Ct ranges associated with infectiousness (Ct <30), >250 hrs of shedding would be required. Thus, our results add weight to an emerging model wherein SARS-CoV-2 airborne emission, at least during tidal breathing, does not implicate FA produced by either acutely symptomatic or hospitalized individuals. Instead, airborne transmission would require aerosol generating procedures, or evaporative shrinking of LDs after exhalation under favourable environmental conditions.

However, both particle emission rates rise and particle size distribution changes with increased volume of speech, as well as coughing (64–67). Here, we show that forced expiration allows for the detection of SARS-CoV-2 in distal lung FA EBC by increasing its concentration. In the patient with highest viral load if FA EBC, Ct’s as low as 25.8 were observed in condensate fractions equivalent to 1 minute’s worth of forced expiration. Since FA EBC viral load also correlated with host 18S rRNA concentration under forced expiration, our results demonstrate that airborne transmission potential increases because forced expiration enhances the nebulisation of airway lining fluid into FA, and not just due to saliva-laden LD evaporative shrinkage in the environment. Thus, daily activities involving forced expiration maneuvers other than symptomatic coughing or medical aerosol generating procedures produce fine aerosols that may feature viral loads high enough to enable transmission of SARS-CoV-2.

## Conclusions

In this study we have evaluated the performance of a new EBC collector design that sought to overcome the key challenges of salivary/ambient contamination, sampling inconsistency and sample loss, and healthcare professional safety (3, 14). This was achieved in a hand-held format by implementing LD impaction ahead of FA condensation in a high thermal capacity condenser, which yields linear sampling of tidal breath FA through vapour nucleation during the inspiration phase of the breath cycle. This physical process promotes highly charged particle aggregation and enlarges FAs to increase capture efficiency by 6x. Resulting FA EBCs have less than 1:1,750 contamination of saliva, are enriched in RNA, contain host metabolites of vesicular nature, and micro-organisms whose viability is a function of condensation temperature. Microbiota analysis of these low biomass samples is achievable, with direct deep sequencing library preparation offering a means of improving upon sample-to-kitome signal problems. However, viral particle disruption cannot be ensured simply through the thermal shock of breath condensation indicating viruses in FA EBCs remain infectious. Use of this device in poorly ventilated clinical wards has confirmed the absence of FA sample contamination from virus-laden environmental aerosols during device use. Efficient SARS-CoV-2 detection among acutely symptomatic COVID-19 cases, however, requires forced expiration which increases distal airway lining fluid concentration in readily airborne, exhaled FA breath fractions.

## Supporting information

Video 2 CFD condensation model

Video 1 PBM-HALE assembly and principles

## Data Availability

All data produced in the present study are available upon reasonable request to the authors

## Author contributions

SAM conceived, and designed in collaboration with IK the new EBC collector, printing early prototypes with the assistance of VT. SA, MC, and SAM performed the production-scale computer aided design and production work and, with the assistance of PH, the computational flow modelling. JH, RG, JAM, TM, SA, EW, WC, LU, BA, AW, KS, and ANel performed all the experimental laboratory work under the supervision and direction of JM, DS, AN, and SAM who designed the experimental work and participated in the data analysis and interpretation. The clinical study was designed by SAM and executed by EJ, NA, PJA, and TN under the supervision of PK, DPK, AT, AK, and MMT. Clinical samples were analysed by AZ, CR, and DCQ under the supervision of GM, PL, RSA, and MMT. JH and SAM co-authored the manuscript. All authors reviewed and agreed the final version of the manuscript.

This project was supported by the InnovateUK iCURE III project no. 43055, and the Northern Accelerator II project no. 25R18P02557. JAM received funding from the German Research Foundation. Airborne particle size analysis was enabled by the kind loan of equipment by Particles Plus Inc.

We thank Professors Aris Katzourakis, Giorgios Sourvinos, Peter J Barnes, and Manfred Frick regarding study societal impact considerations, organisational aspects of conducting sampling at PAGNI Hospital, the observation of no *Haemophilus* among FA microbiota, and with manuscript review, respectively.

---

**Figure S1:**
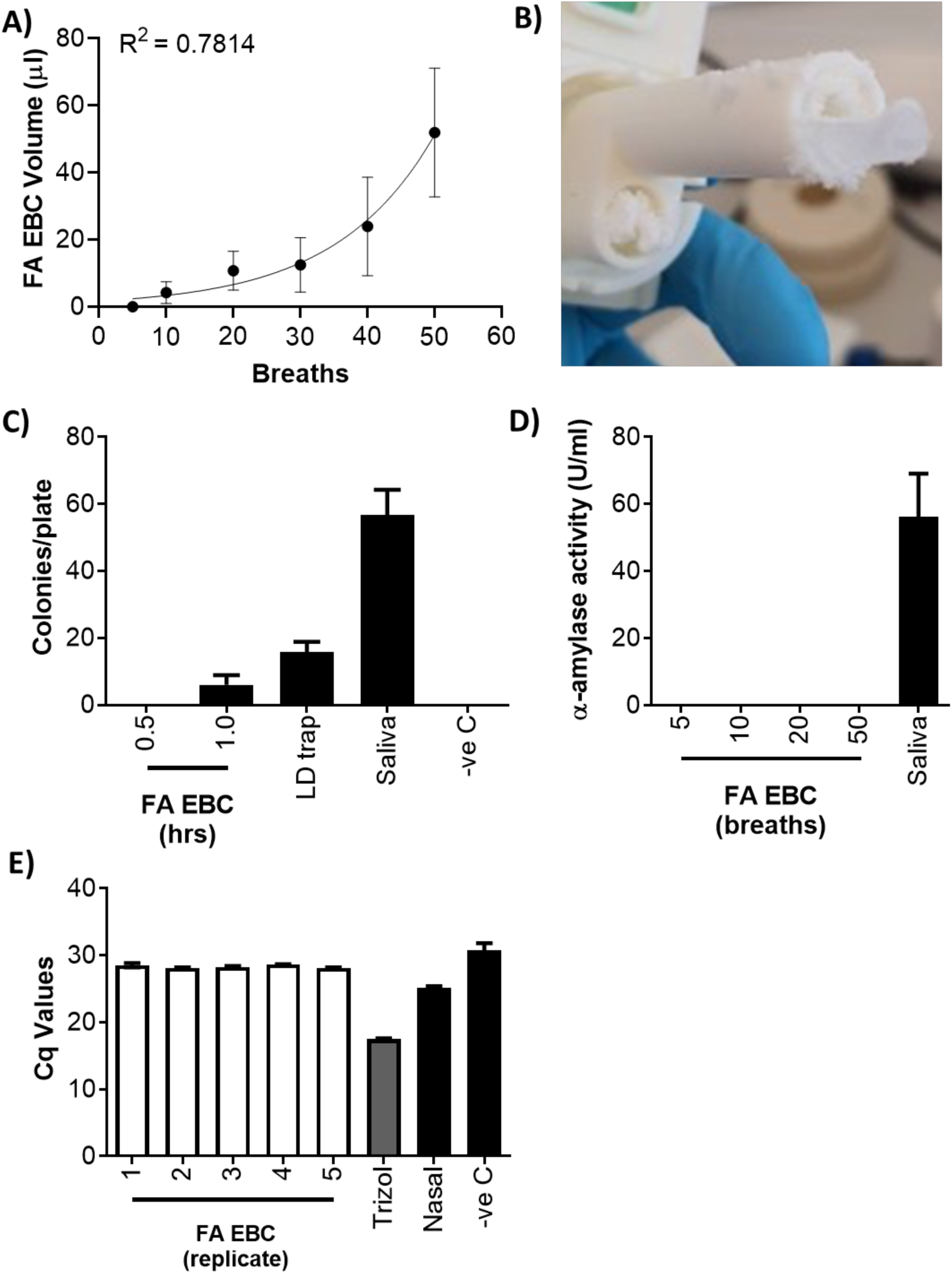
Impact of PBM-HALE^TM^ design optimization. (A) FA EBC sampling reliability in early 3D printed prototypes (n=5; averages ± standard deviation shown; R^2^ value for a log_10_-linear regression analysis shown). (B) Dreschel tube wall architecture effects on platform lid thermal properties resulting in condensation within the Dreschel tubes when using −78.5°C condensation (C) Effect of continuous sampling in FA EBC microbial viability as determined by blood agar plating of adult healthy male volunteer samples obtained using −78.5°C condensation (n=5; averages ± standard deviations shown). (D) Salivary vs FA EBC sample α-amylase activity in an adult healthy male volunteer using −100°C condensation (n=5; averages ± standard deviation). (E) Pan-eukaryotic 18S rRNA levels in 5 independent, 20 tidal breath samples (EBC 1-5), a 30min sample (EBC 6), and a nasal swab sample obtained from an adult healthy male volunteer using −78.5°C condensation (averages ± standard deviations of triplicate technical RT-qPCR replicates shown).

**Figure S2:**
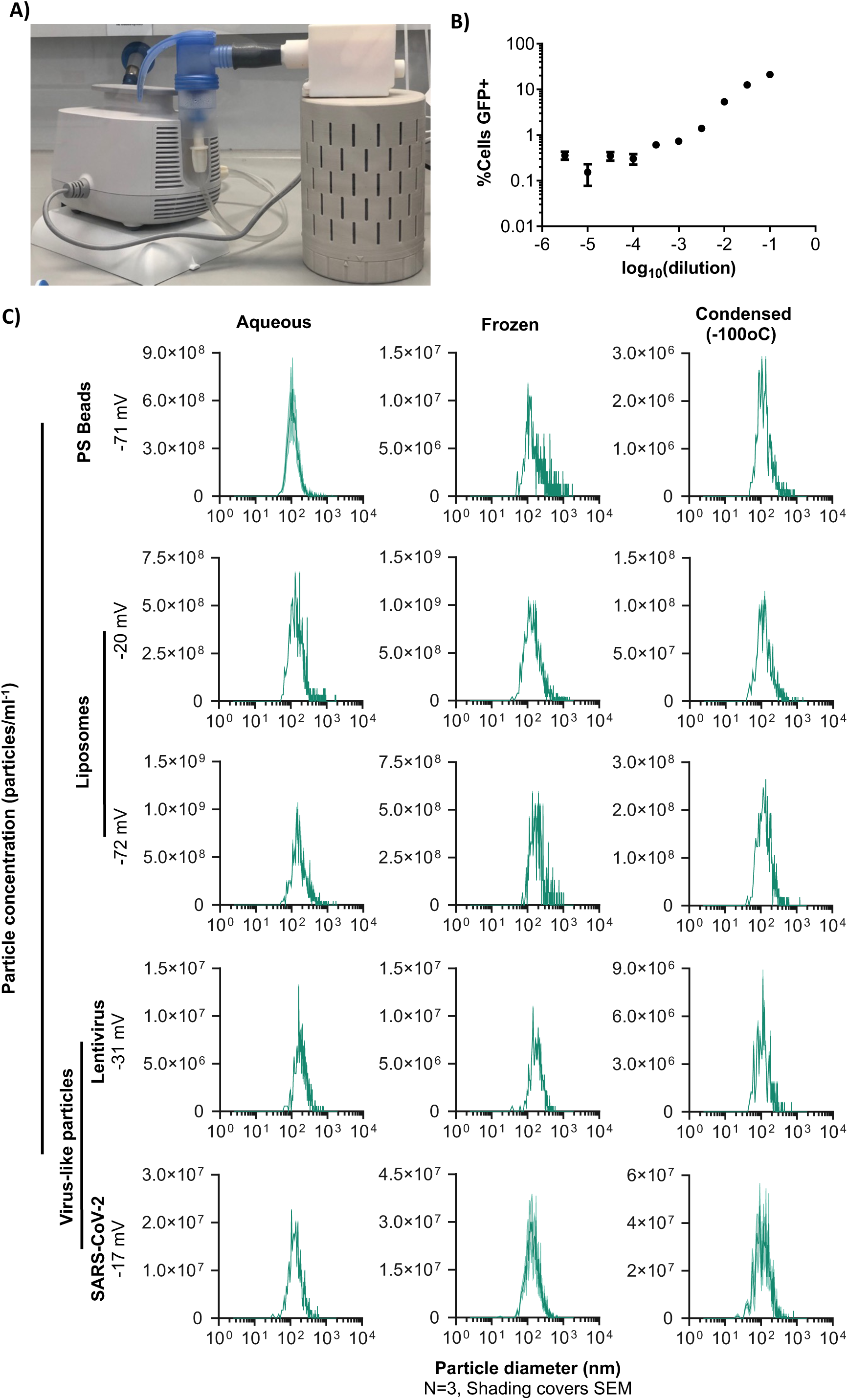
Impact of condensation temperature on mechanically nebulized infectious pseudoviruses and synthetic nanoparticles. (A) A clinical grade nebulizer (PARI) was attached to PBM-HALE^TM^ using a 3D printed adapter to passively route suspensions of nanoparticles into the condenser. (B) Titration of VSV-GFP lentivirus for the determination of TCID50 identifies a 2.5 log stock dilution as the appropriate concentration for the above-background detection of GFP positive cells by fCM (average ± standard deviation of 3 independent replicates). (C) Effect of flash-freezing or mechanical aerosolization/condensation capture of polysterene beads (PS Beads), liposomes, lentiviral virus-like particles (Lenti), or SARS-CoV-2 virus-like particles on particle concentration as a function of particle diameter, as measured by nanoparticle tracking analysis (average ± standard error of the mean of 3 independent replicates is depicted in the green-shaded area). The charge (zeta potential) of each particle type is shown.

